# Association of current *Schistosoma mansoni, S. japonicum,* and *S. mekongi* infection status and intensity with periportal fibrosis: a systematic review and meta-analysis

**DOI:** 10.1101/2024.04.09.24305558

**Authors:** Adanna Ewuzie, Lauren Wilburn, Dixa B. Thakrar, Nia Roberts, Reem Malouf, Goylette F. Chami

## Abstract

**Background:** Periportal fibrosis (PPF) is a severe morbidity caused by both current and past exposure to intestinal schistosomes. We assessed the association between current/active infection status and intensity of *Schistosoma mansoni, S. japonicum,* or *S. mekongi* with PPF.

**Methods:** We systematically searched the Cochrane Central Register of Controlled Trials, Embase, Global Health, Global Index Medicus and Medline on August 24, 2022. A meta-analysis to derive pooled effect sizes for concurrently measured current schistosome infection status and intensity against author-defined PPF was conducted using inverse-variance weighted random effects. Subgroup analyses by study characteristics and risk of bias assessments using a modified National Institute of Health Risk of Bias Tool were completed. The protocol was prospectively registered on PROSPERO (CRD42022333919).

**Findings:** We identified 2646 records; 37 studies were included in the systematic review and 30 studies in the meta-analysis. *S. mansoni* was most studied (91·89%; 34/37). PPF was heterogeneously defined with the Niamey ultrasound protocol commonly used for diagnosis. Individuals with any current infection were 2·50 (95% CI:1·71-3·66) times more likely to have PPF compared to uninfected individuals with high heterogeneity (I^2^ statistic 94·80%). Subgroup analyses showed there was no association when only ultrasound patterns or modified Niamey Protocols were used. There was no association in studies conducted in sub-Saharan Africa after 2002 when mass drug administration became widespread, or in studies with a low risk of bias. No significant association was found between schistosome infection intensity and PPF.

**Interpretation:** World Health Organization guidelines use current schistosome infection intensity as a proxy for schistosomiasis-related morbidity. This study supports that only current infection status was tenuously associated with PPF. Guidelines are needed to better monitor schistosomiasis-related morbidities.

**Funding:** NDPH Pump Priming Fund, Wellcome Trust-ISSF (204826/Z/16/Z), John Fell Fund, Robertson Foundation, and UKRI EPSRC (EP/X021793/1).

**Research in Context:** *Evidence before this study:* Periportal fibrosis (PPF) is a severe complication of intestinal schistosomiasis. We searched the Cochrane Central Register of Controlled Trials, Embase, Global Health, Global Index Medicus, and Medline from the database inception to August 24, 2022. The broad search terms were “Schistosoma”, “fibrosis” AND “periportal OR liver”. Three reviews were detected by the search string; these detailed how human genetics influence fibrosis outcomes, non-invasive methods of periportal fibrosis in schistosomiasis patients, and human host regulation of liver fibrosis during schistosome infection. Outside this search string, reviews exploring the impact of co-infections on liver morbidity (Hepatitis B/C and malaria), the use of ultrasonography for assessing morbidity, and the impact of chemotherapy on liver morbidity were identified or in progress. No review had assessed the impact of current intestinal schistosome infection status or intensity on PPF outcomes.

*Added value of this study:* Here we provide quantitative evidence for the influence of (or lack thereof) current *Schistosoma mansoni, S. japonicum, and S. mekongi* infection status and intensity on PPF while presenting the risk of bias in the available literature. By synthesising data ranging from 1988–2020 encompassing 17317 participants, across all age ranges, we found that individuals with current schistosome infection were 2·50 times more likely to have PPF when compared to individuals who are not currently infected. Heterogeneity was high (>90%) across studies and was not reduced when moderate or high risk of bias studies were excluded. The association of current schistosome infection status was tenuous, determined solely by unadjusted studies that ignored cofounders and were conducted prior to mass drug administration. The association was observed only in moderate to high risk of bias studies and not present in low risk of bias studies. Importantly, we found no significant association between the intensity of current schistosome infections and PPF with very few studies available on current infection intensity.

*Implications of all the available evidence:* Current World Health Organization (WHO) guidelines focus on reducing schistosomiasis-related morbidity as approximated by community prevalence cut-offs set based on only current schistosome infection intensity. This meta-analysis provides evidence that those currently infected with schistosomes had an increased likelihood of having PPF, but only when infection status was considered rather than infection intensity. The high heterogeneity found among studies presented here suggests the need for standardisation of PPF diagnosis to accurately estimate the global burden of this disease in the future. Our findings suggest that in the current context of widespread, repeated mass drug administration infection proxy indicators are poor estimates of severe morbidity related to schistosomal liver fibrosis. Guidelines or recommendations are needed now from the WHO to assist endemic countries on how to directly monitor schistosomiasis-related morbidities as opposed to monitoring current infections while considering existing local resources and health system constraints.

## INTRODUCTION

Periportal fibrosis (PPF) is a severe but common complication of the intestinal forms of the parasitic disease, schistosomiasis. PPF disproportionately affects the poorest communities in Africa.^1^ Transmission of the causative trematode of the species *Schistosoma mansoni, S. japonicum, or S. mekongi* requires human contact with freshwater that is contaminated with parasite eggs, in addition to the presence of competent snail species as the intermediate host.^2^ Once individuals become infected the flukes migrate to the liver then once male-female paired and matured, move to reside in mesenteric venules. Here, female flukes will produce 100-3000 eggs per day, depending on the species.^3^ Not all eggs are excreted and can be swept back into the liver and spleen where granulomatous inflammation ensues, which may lead to PPF. PPF is a type of fibrosis distinguished by distinct patterns around the hepatic portal tracts of the liver.^2,4^ Ongoing fibrosis causes damage and eventual blockages of the liver vasculature forcing local restructuring.^5^ Left untreated, the disease can cause portal hypertension, upper gastrointestinal bleeding, and ultimately premature death.^2^

Mass drug administration (MDA) of praziquantel is recommended by the World Health Organisation (WHO) to control schistosomiasis infection and morbidity.^6^ The availability of MDA has successfully led to widescale, significant reductions in *Schistosoma* infection prevalence and infection intensity.^6^ Yet, PPF and related morbidities remain in treated communities despite repeated rounds of MDA.^7^ A meta-analysis by Andrade et al. shows MDA better reduces morbidity related to urinary forms rather than intestinal forms of schistosomiasis.^8^

The current WHO guidelines recommend administering praziquantel annually to individuals over the age of two years in communities with an infection prevalence >10%.^6^ However, estimated infection prevalence is heavily skewed by school-aged children, who typically have the highest prevalence and intensity of infection but low levels of PPF.^9^ PPF is associated with chronic exposure to schistosomes, and the time taken from the initial infection to the development of severe fibrosis may range from 5-15 years.^2^ The time lag for PPF development results in scenarios where treatment guidelines may not align with morbidity prevalence by age group.^7^ When coupled with the risk of reinfection, the contribution of chronic/historical infections, and confounders of other co-infections or alcohol use, it remains unclear how current infection status relates to PPF.^2^ In the context of repeated MDA, individuals have varying histories of treatment, so heterogeneity may be introduced into what is known about the relationship between current schistosome infection and PPF.^10^ Despite this uncertainty, WHO guidelines suggest that <1% prevalence of heavy infection intensities (<400 eggs per gram of stool) in school-aged children approximates the elimination of morbidity as a public health problem for schistosomiasis.^6^

Here, we quantify the pooled effect size for current schistosome infection status and intensity with PPF status in schistosomiasis-endemic regions.

## METHODS

### Search strategy and selection criteria

In this systematic review and meta-analysis, we searched the Cochrane Central Register of Controlled Trials(Cochrane Library, Wiley)[Issue 7 of 12, July 2022], Embase(OvidSP)[1974-], Global Health(OvidSP) [1973 to 2022 Week 19], Global Index Medicus(https://pesquisa.bvsalud.org/gim/) and Medline(OvidSP)[1946-] from database inception to August 24, 2022. The broad search terms covered title, abstract, author keywords and subject headings for “Schistosoma”, “fibrosis” AND “periportal OR liver”. We applied methodological filters to limit to RCTs or observational studies.^11,12^ Animal studies were excluded, no date or language limits were applied, full details are in appendix 1, pp 2-4. References were exported to Covidence for screening and deduplication.^13^ The study protocol was prospectively registered on PROSPERO (CRD42022333919) on May 19, 2022, and is reported according to Preferred Reporting Items for Systematic reviews and Meta-Analyses (PRISMA) guidelines.^14^

We excluded non-human and non-English studies, reviews, editorials, personal opinions, and case reports. Self-reported infection status was an ineligible exposure. PECO inclusion criteria are described in detail in appendix 1, p5. Two reviewers (AE, DT) independently screened abstracts and full-text reports for eligibility. A third reviewer (RM) was consulted in the case of disagreement. AE and LW extracted the data and DT independently cross-checked 20% of the data extractions.

The outcome of periportal fibrosis was recorded as defined by the study authors. For the key exposure of current/active infection, data was extracted for *Schistosoma* species, diagnostic, and the author-provided infection status and intensity definitions. Additional information such as study design and setting, age and sex of participants, and ultrasound protocol also were recorded (appendix 1, p6). If effect sizes were not provided by study authors, odds ratios were reconstructed where available from in-text information or as provided by authors when contacted for more information.

### Data analysis

Random effects models using the generic inverse variance method were used to calculate pooled effect measures (ORs) and the 95% CI in Stata/MP v17. Overall heterogeneity was calculated using Higgin’s I^2^ statistic to explain between-study variation.^15^ Subgroup analyses were conducted when at least three or more studies were available. Subgroups considered were continent, *Schistosoma* species, study setting, before/after MDA in sub-Saharan Africa, ultrasound protocol, PPF outcomes definitions, and Niamey Deviations (appendix 1, p7). The risk of bias (ROB) was assessed using a modified National Institute of Health Risk of Bias Tool from the National Heart, Lung, and Blood Institute (appendix 1, pp8-10). Publication bias was evaluated using funnel plots and Egger’s regression.^16,17^

## RESULTS

The PRISMA flow chart for the search results is presented in Figure 1. The electronic search retrieved 2646 records; and 908 duplicate records were removed at this stage. 1729 abstracts were screened and records that did not meet the inclusion criteria were removed. A total of 268 articles were screened at the full-text stage (appendix 1, pp11-25). 37 studies were eligible for the systematic review and 30 studies were included in the meta-analysis.

**Figure 1:**
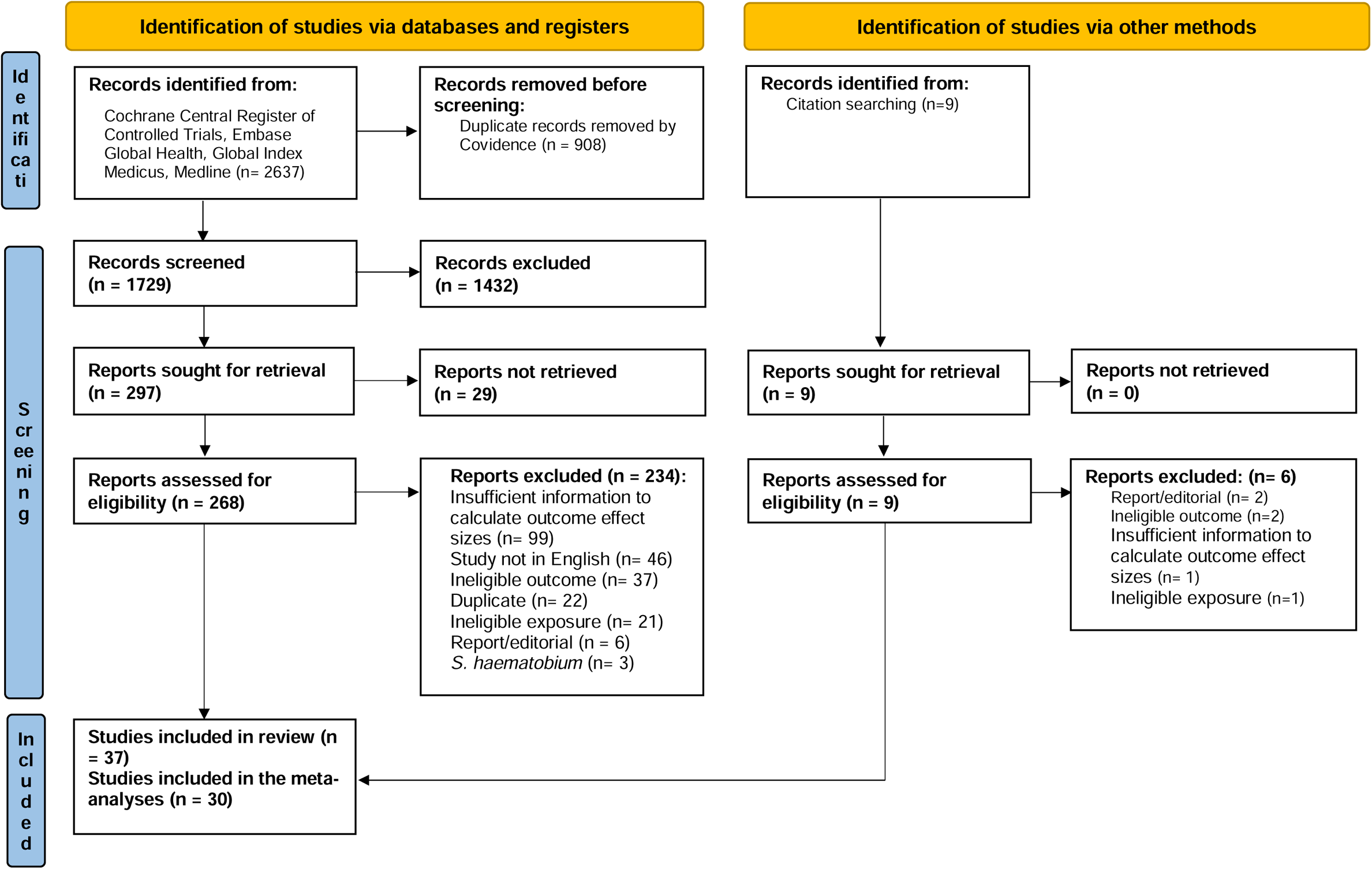
PRISMA study selection.

Study characteristics are summarised in Table 1, full details are provided in Table 2. Across 37 studies^18–54^, there were 31295 participants. Egypt was the most frequently studied country ((Figure 2) (21**·**62%; 8/37))^18,20,21,26,32,36,42,43^. Most studies included both men and women (97**·**29%, 36/37), and a single study included only men (2**·**70%, 1/37).^39^ Age categories were inconsistently reported throughout the studies without clear minimum or maximum ages. Some studies diagnosed other infections including malaria (10·81%, 4/37),^23,28,30,33^ *S. haematobium* (5·40%, 2/37),^25,34^ HIV (5**·**40%, 2/37),^33,44^ soil-transmitted helminths 5**·**40%, 2/37),^38,40^ hepatitis B/C co-infection (8·10%, 3/37),^20,22,30^ and *Opisthorcis viverrini* (a species of liver fluke) (2**·**70%, 1/37).^50^ However, many did not provide data on the co-infection with *Schistosoma* species (72**·**72%, 8/11). Most studies diagnosed schistosome infection by Kato-Katz microscopy (91**·**89%, 34/37)^18–25,28–47,49–52,54^ with two studies using only serology^27,44^ and three studies with both microscopy and serology.^26,48,53^ Over 43% (16/37)^19,22,23,30–34,37,38,41,44,49,52,54^ of all studies utilised the Niamey protocol and almost 30% used the Cairo protocol (11/37).^18,20,21,25,36,42,43,45,48,51,53^ Eight reports used non-WHO approved ultrasound protocols (21**·**62%, 8/37)^24,26–29,35,39,46^ and three studies did not report the protocol that was used (8**·**10%, 3/37).^40,47,50^

**Figure 2:**
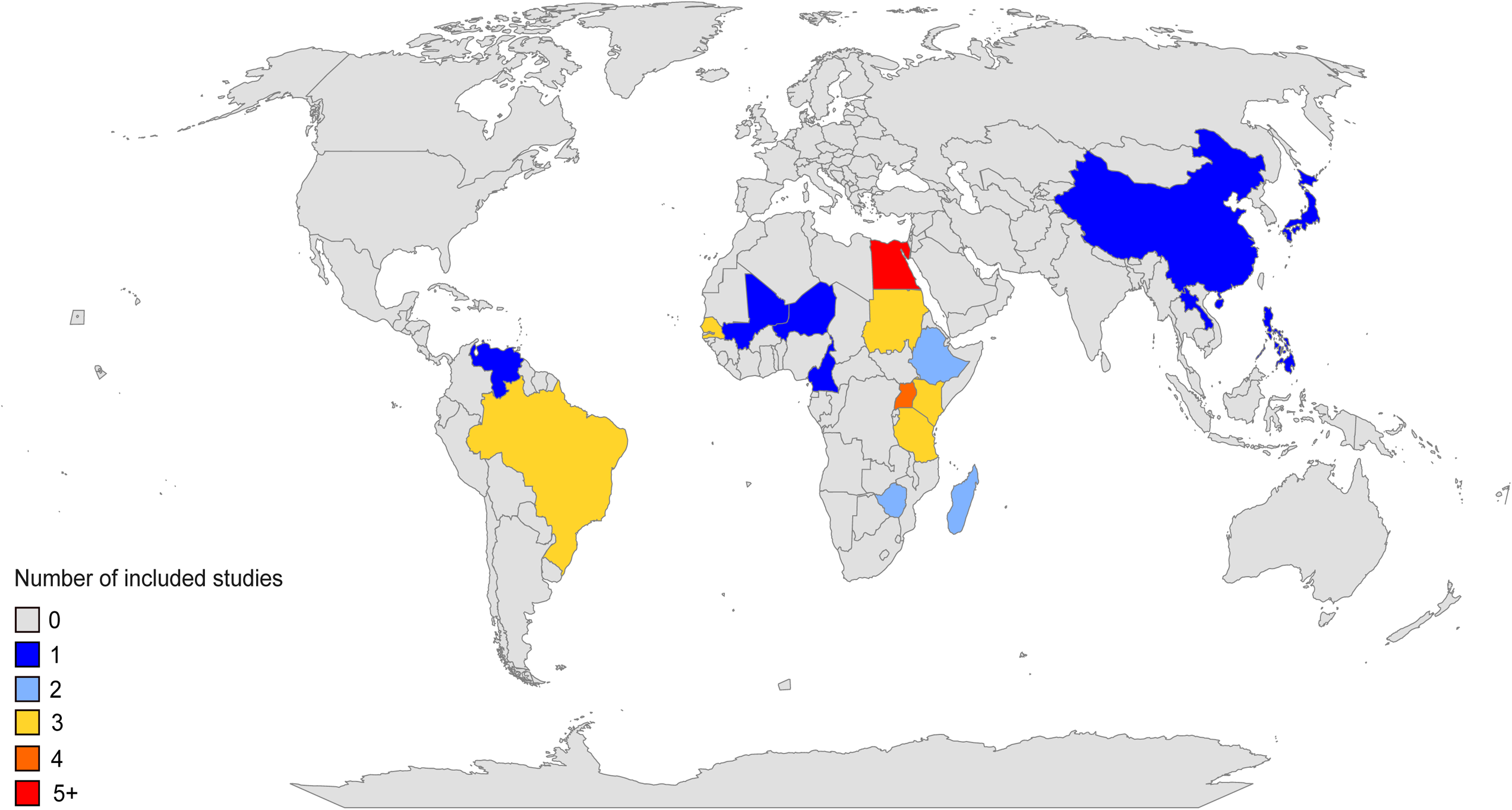
Map showing locations and number of studies included in systematic review.

**Table 1:** Summary of study characteristics included in the systematic review. The total number of studies included here is 38 as King et al, 2003^37^ included two separate study populations, from different countries.

| Study characteristic | Type | Number of participants | Number of studies | Percentage (%) |
| --- | --- | --- | --- | --- |
| Study Aims | Prevalence of schistosome infection and/or morbidity | 9635 | 14 | 36.84 |
|  | Asses the use of ultrasound in the diagnosis of fibrosis. | 9405 | 9 | 23.68 |
|  | Immune/genetics of periportal fibrosis | 2915 | 6 | 15.78 |
|  | Association of schistosome infection and periportal fibrosis | 941 | 3 | 7.89 |
|  | Impact of praziquantel on schistosomiasis morbidity | 1838 | 2 | 5.26 |
|  | Risk factors in transmission | 611 | 2 | 5.26 |
|  | Disease progression | 792 | 1 | 2.63 |
|  | Impacts of infection intensity | 5158 | 1 | 2.63 |
| Study region | Sub-Saharan Africa | 18110 | 19 | 50.00 |
|  | North Africa/East | 10960 | 11 | 28.95 |
|  | South-East Asia | 808 | 2 | 5.26 |
|  | Central Asia | 700 | 2 | 5.26 |
|  | South and Central America | 717 | 4 | 10.53 |
| Study design | Cross-sectional | 21885 | 29 | 76.32 |
|  | Cohort | 7009 | 3 | 7.89 |
|  | Case-control | 2401 | 6 | 15.79 |
| Study Setting | Community | 23070 | 26 | 68.42 |
|  | School | 6714 | 5 | 13.16 |
|  | Health clinic/hospital | 833 | 5 | 13.16 |
|  | Workplace | 435 | 1 | 2.63 |
|  | Community + Health Clinic | 243 | 1 | 2.63 |
| *Schistosoma* species | *S. mansoni* | 29787 | 34 | 89.47 |
|  | *S. japonicum* | 1265 | 3 | 7.89 |
|  | *S. mekongi* | 243 | 1 | 2.63 |
| *Schistosome*<br>diagnostic tool | Kato-Katz Microscopy | 30612 | 35 | 92.11 |
|  | Serology | 122 | 1 | 2.63 |
|  | Antigen | 299 | 1 | 2.63 |
|  | Microscopy and Serology | 262 | 1 | 2.63 |
| Ultrasound protocol | Niamey | 8557 | 16 | 42.11 |
|  | Cairo | 12528 | 11 | 28.95 |
|  | Abdel-Wahab | 1984 | 2 | 5.26 |
|  | Uto | 2092 | 1 | 2.63 |
|  | Homeida | 1113 | 2 | 5.26 |
|  | Managil | 1671 | 1 | 2.63 |
|  | Doehring-Schwerdtfeger | 600 | 1 | 2.63 |
|  | Undefined | 1677 | 1 | 2.63 |
|  | Not reported | 1073 | 3 | 7.89 |
| Author defined<br>periportal fibrosis<br>definition | Mean of three measurements | 8185 | 9 | 23.68 |
|  | Image patterns C-F | 10893 | 13 | 34.21 |
|  | Grade based on echogenicity | 3405 | 5 | 13.15 |
|  | Not reported | 5548 | 2 | 5.26 |
|  | Other | 3240 | 9 | 23.68 |

**Table 2.**
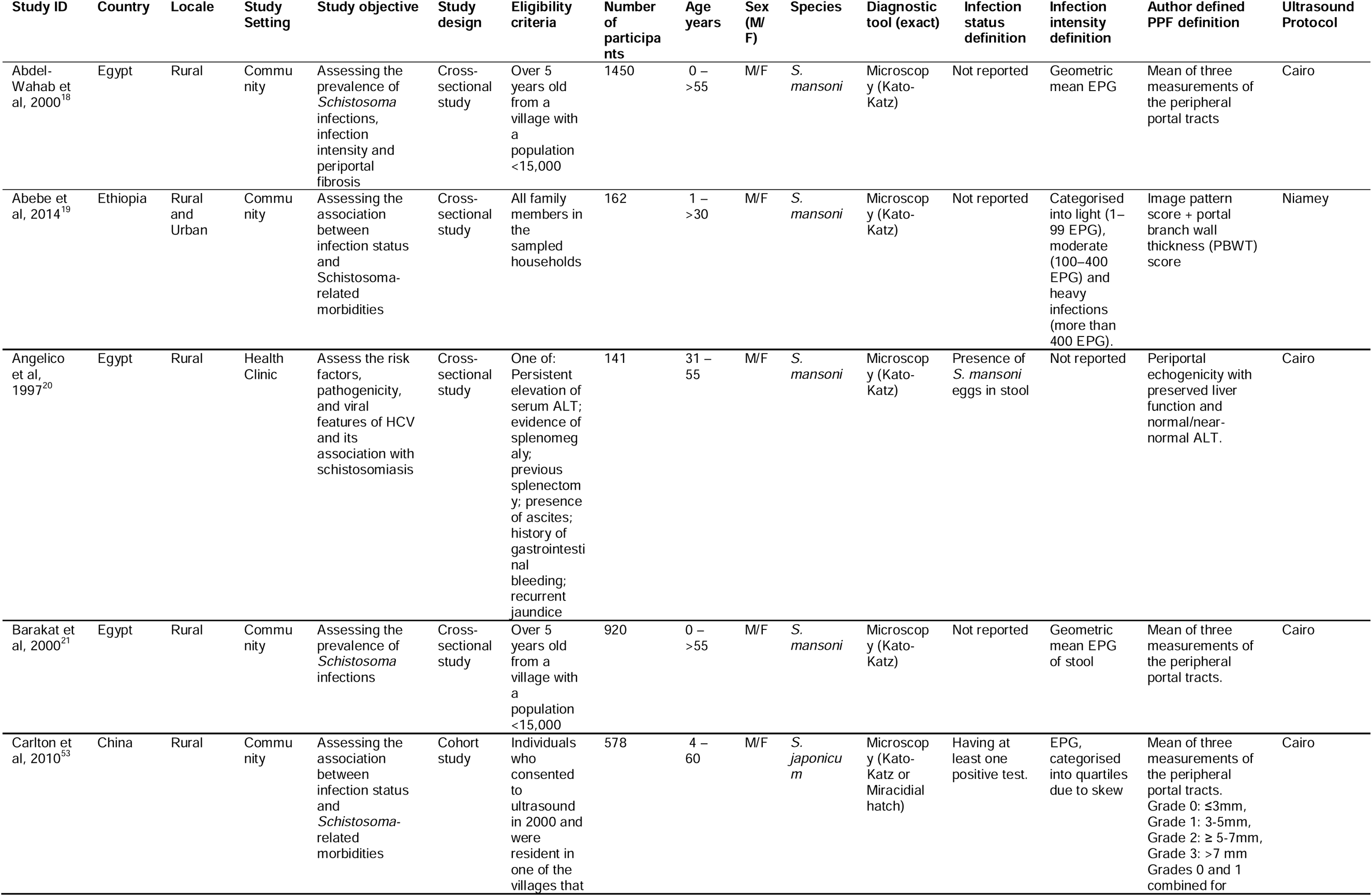
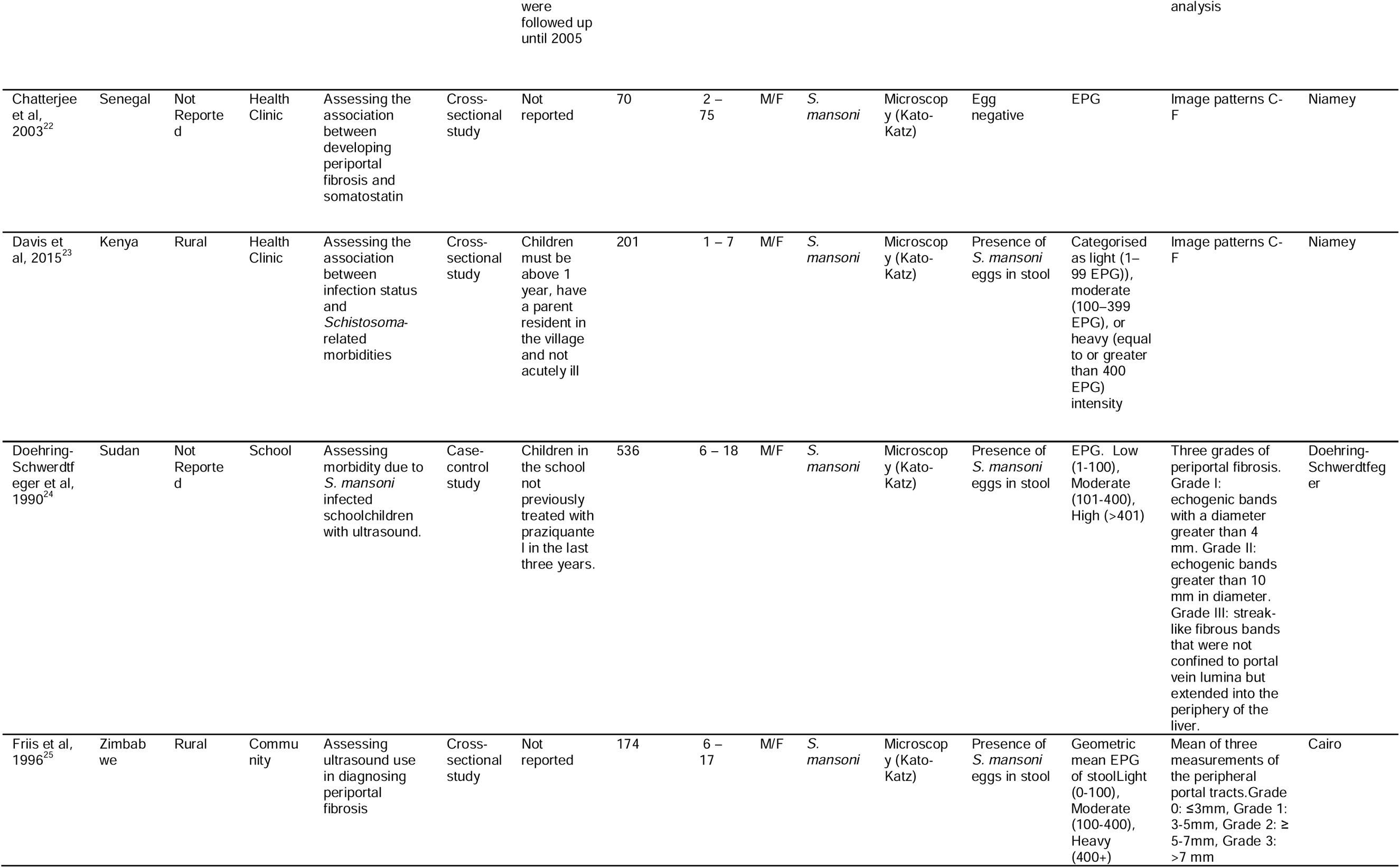
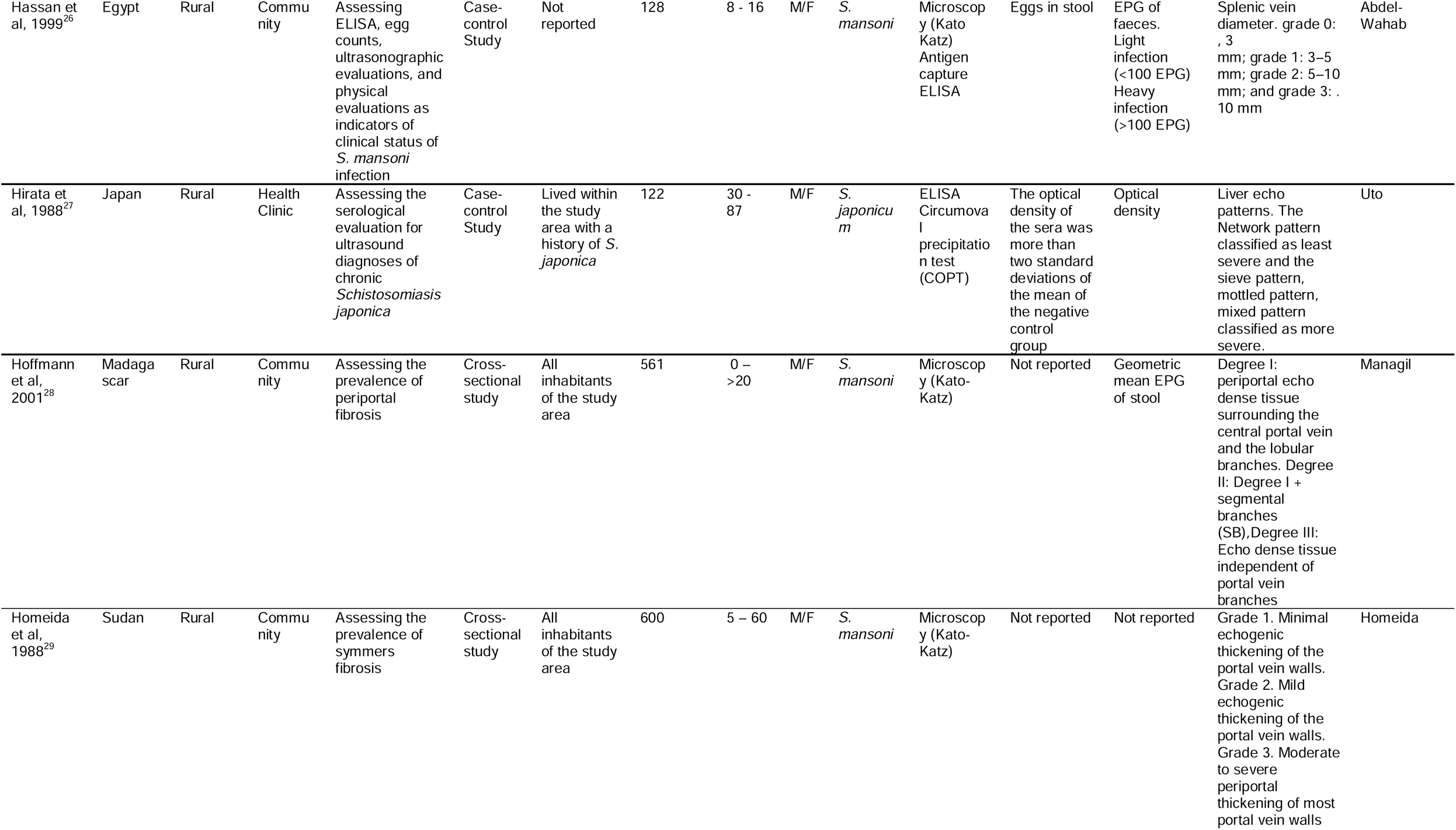
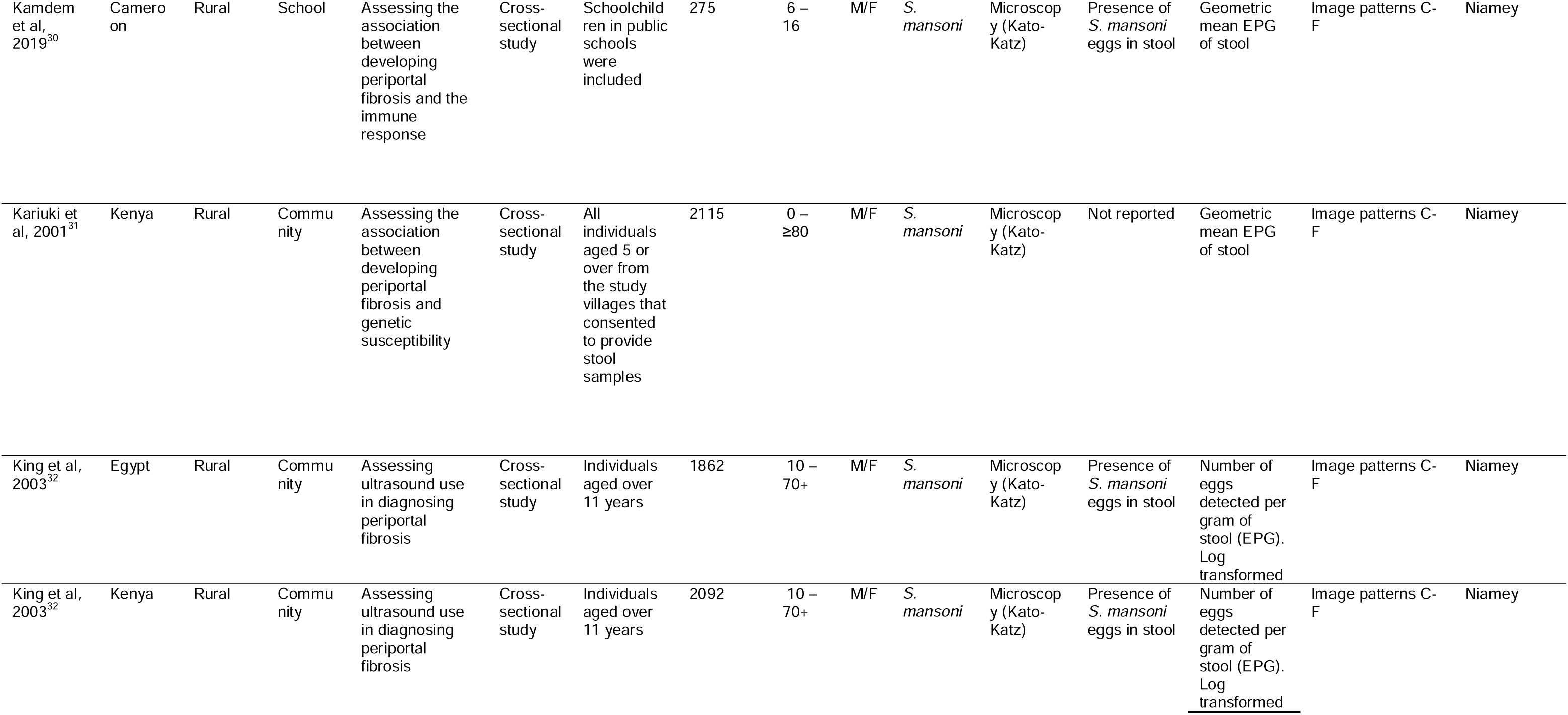
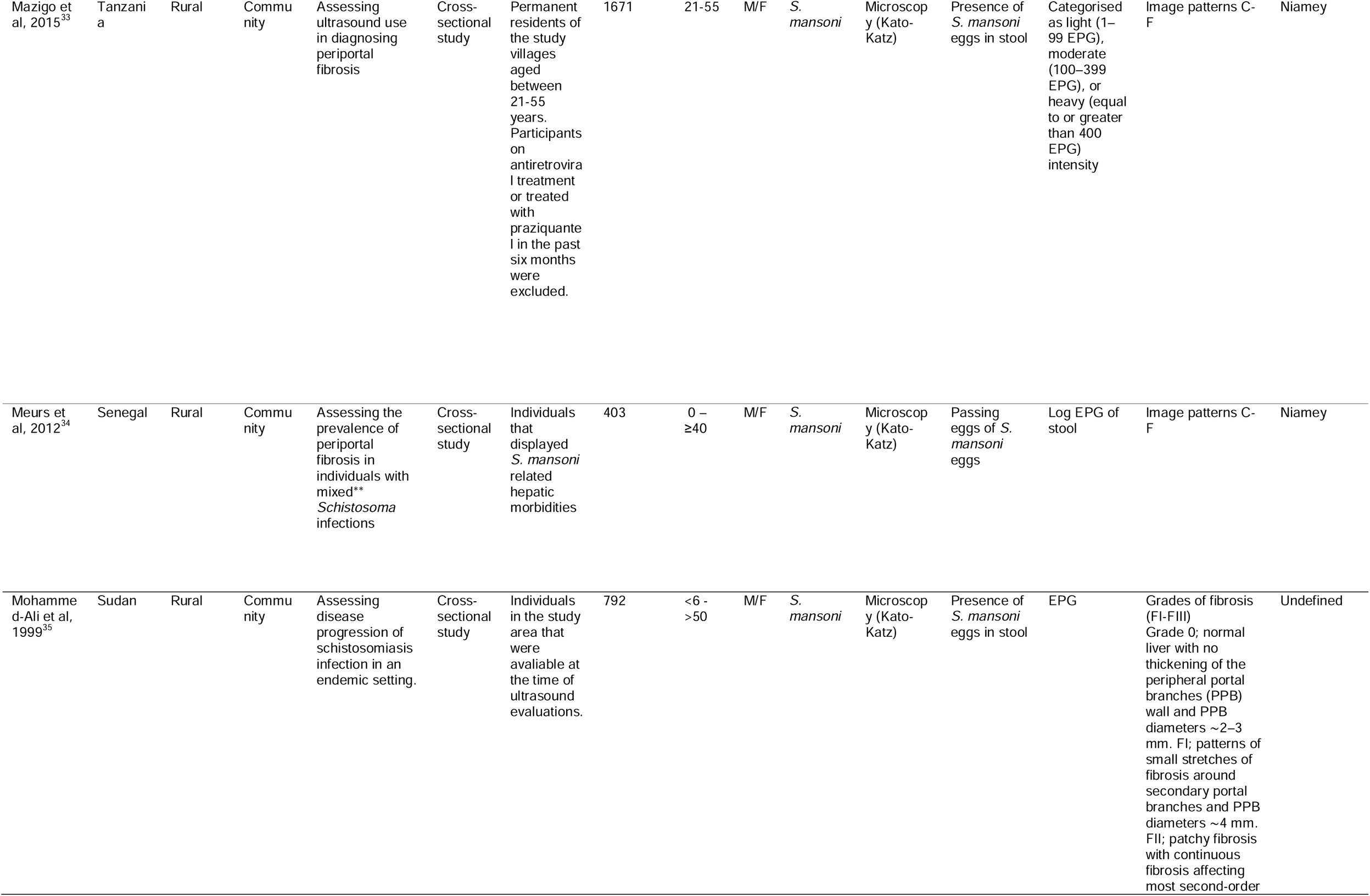
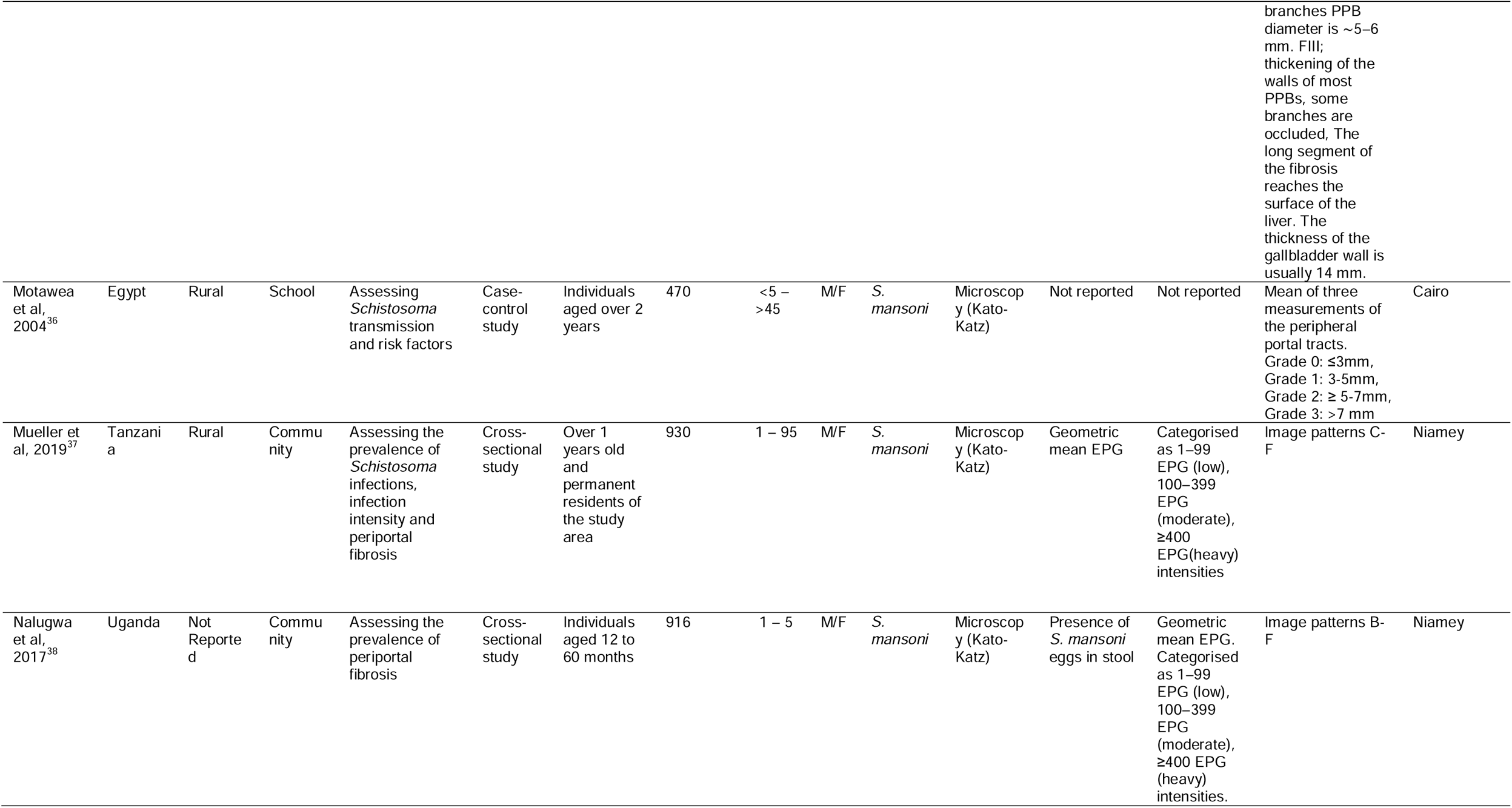
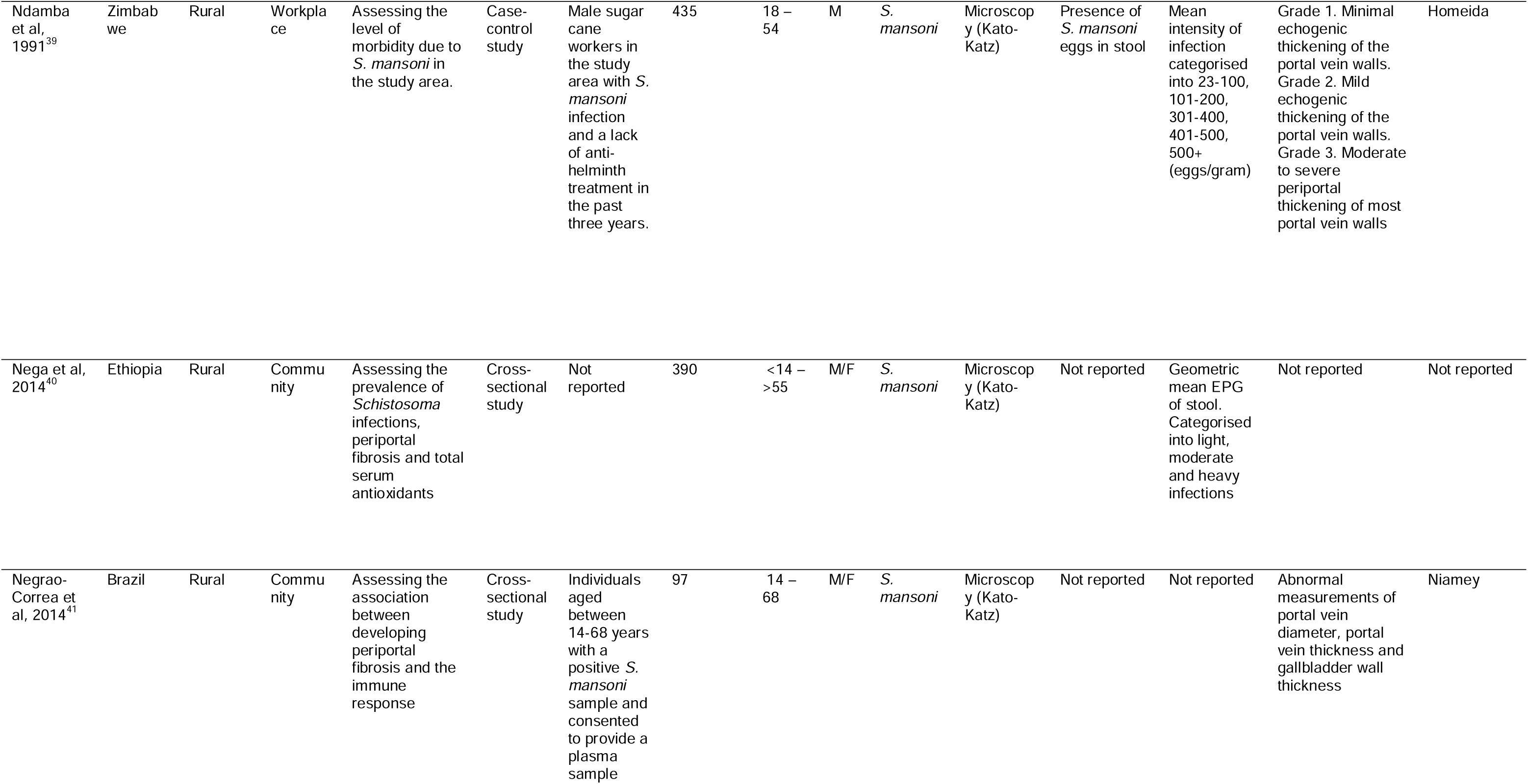
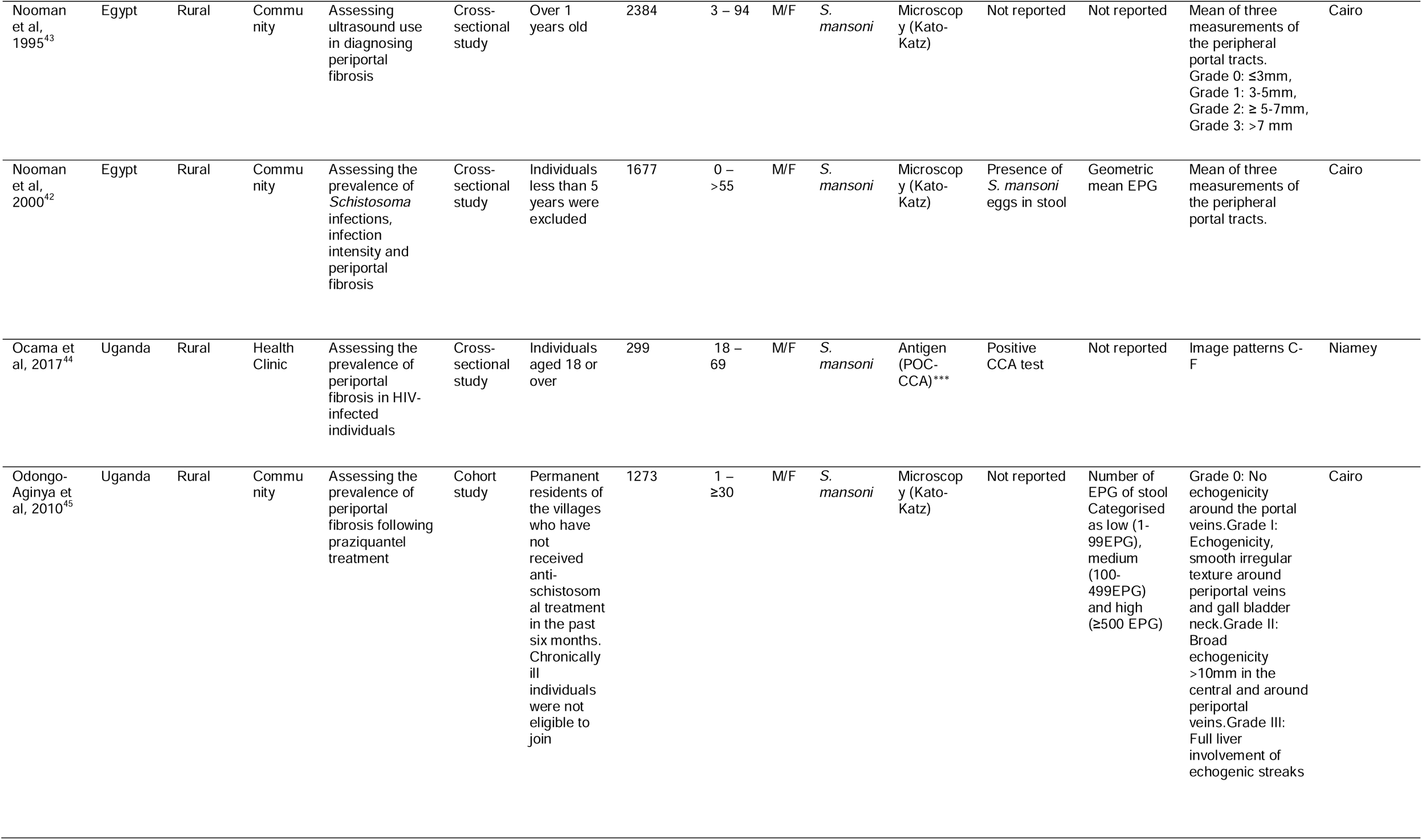
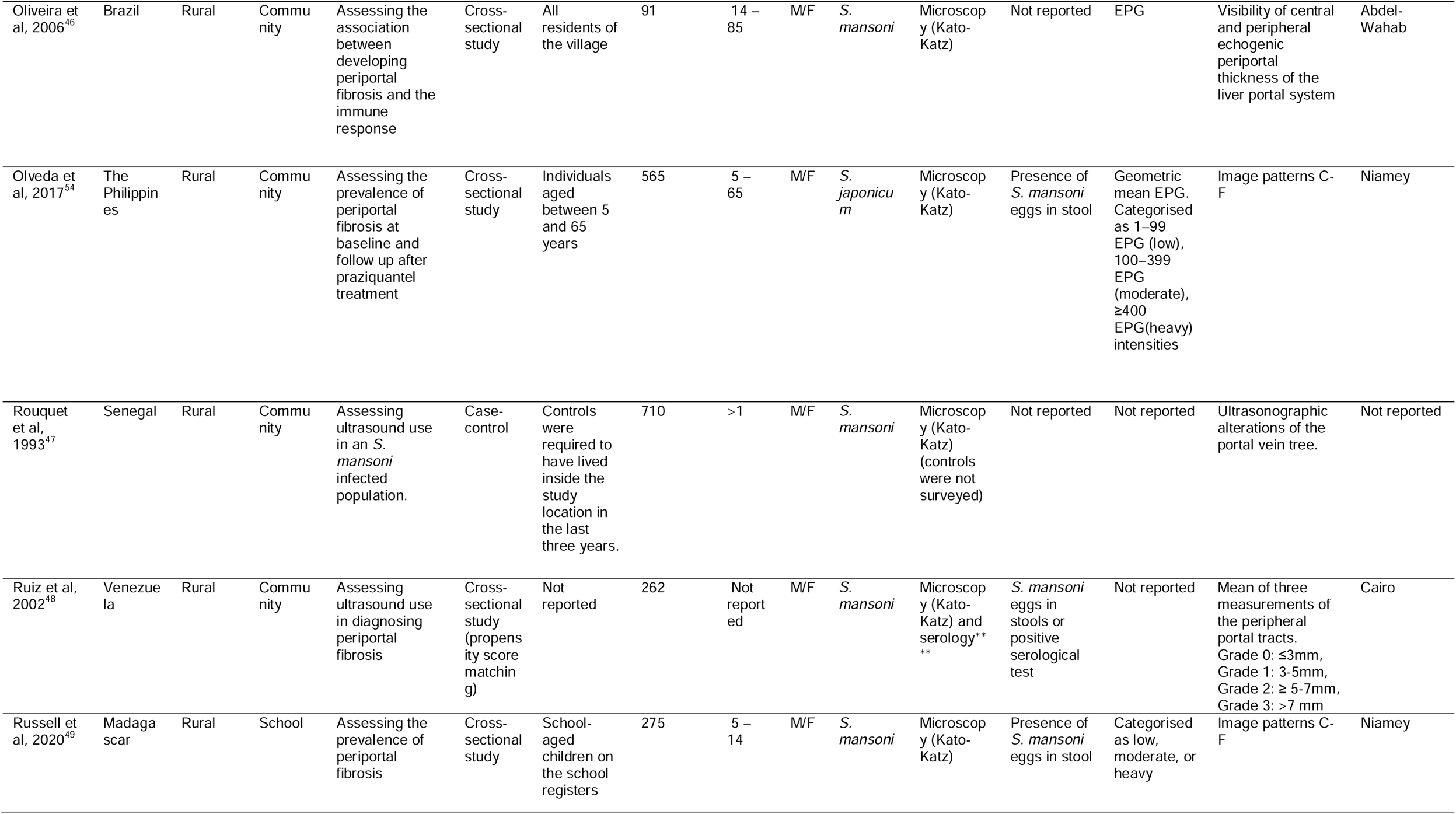
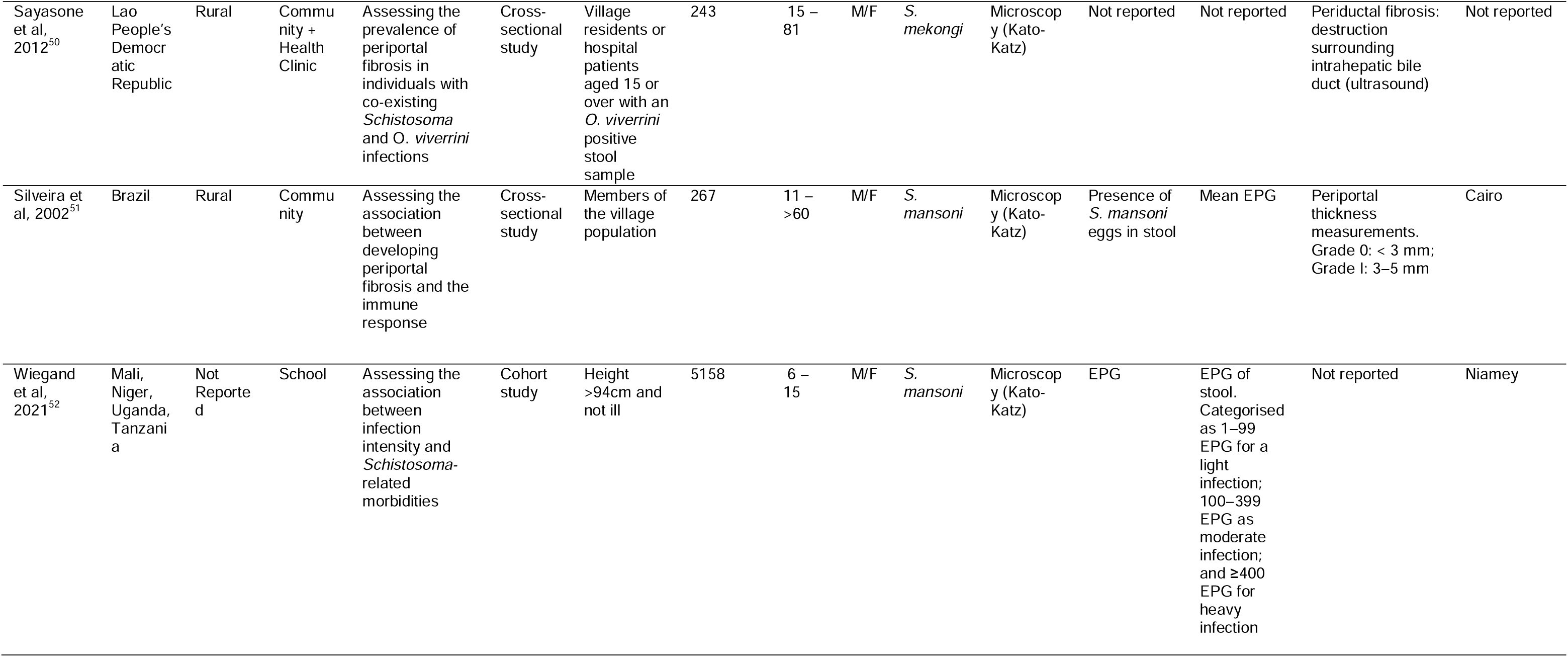
Study Characteristics. * Number of people measured for both the exposure and the outcome **Mixed referring to co-infection with *S. mansoni* and *S. haematobium* *** Specific test(s): Rapid Medical Diagnostics, Pretoria, RSA **** 54.9% of the sample was diagnosed using microscopy, 45.1% of the sample was diagnosed by serology. Study ID: author and date of publication ALT: Alanine transaminase COPT: Circumoval Precipitin Test and Alkaline Phosphatase Immunoassay ELISA: enzyme-linked immunosorbent assay. EPG: Eggs per gram of stool HCV: Hepatitis C

Thirty studies, involving 17317 participants were eligible for inclusion in the meta-analysis. See appendix 2 for the full data extraction table. Seven studies were not included in the meta-analysis as the data presented could not be combined with at least two other studies. For example, studies presenting summary data on the mean eggs per gram without separating by intensity category. When findings were pooled, current schistosome infection status was associated with 2**·**50 times higher likelihood PPF when compared to no current infection ((OR 2**·**50, 95% CI: 1·71-3·66, p<00·1) (Figure 2)). This pooled effect had high heterogeneity (I^2^=94**·**80%). Three studies reported over 30 times greater odds of having PPF when infected with schistosomes with even larger confidence intervals.^24,26,47^ Although these individual effect sizes were high, removing them did not reduce the overall heterogeneity (appendix 1, p26).

Current schistosome infection status and its effects on PPF were mostly extracted from unadjusted analyses without any consideration for confounders. A single study (1/30) presented adjusted effect measures for the association between current schistosome infection status and PPF, however, this could not be included in the subgroup analyses due to a lack of at least two other studies as comparators.^23^ Here the unadjusted prevalence ratio was 1·60 (95% CI: 0·60-4·50) and after adjustments for sex and age, the effect size was slightly reduced to 1·50 (95% CI: 0·50-9·0). Four additional studies provided adjusted effect sizes for risk factors of PPF, including age and gender, but did not provide a breakdown of the schistosome infection status, therefore effect sizes could not be extracted.^33,37,49,53^

Subgroup analyses are shown in Table 3. Studies conducted within Asia (OR 2·23, 95% CI: 1·51-3·30) or within Africa (OR 2·23, 95% CI:1·51-3·30) showed an association between current infection status and PPF with no significant associations in South America (OR 1·32, 95% CI:0·60-2·92). *S. japonicum* or *S. mekongi* (OR 5·35, 95% CI:1·13-25·40) species had a greater pooled effect size, but much higher variability when compared to studies with of *S. mansoni* (OR 2·27, 95% CI:1·56-3·32). Within community-based studies (OR 2·27, 95% CI:1·47-3·62) a positive association was observed but no association was present within school-based studies (OR 3·39, 95% CI:0·81-14·13) or health clinics (OR 1·86, 95% CI:0·81-4·31). Case-control studies (OR 15·80, 95% CI:5·70-43·82) presented stronger associations of current infection status with PPF when compared to cross-sectional studies (OR 1·83, 95% CI:1·34-2·49); however, with outlier effect sizes removed, there were not enough studies to meta-analyse. Importantly, in sub-Saharan Africa, before the introduction of MDA in 2003 there was a significant association between current infection status and PPF (OR 5·91, 95% CI:2·29-15·21) but this association was removed after the introduction of MDA in 2003 (OR 1·25, 95% CI:0·82-1·92). Subgroup analyses remained robust when studies showing outlier effect sizes ≥30 were excluded (appendix 1, p27).

**Table 3:**
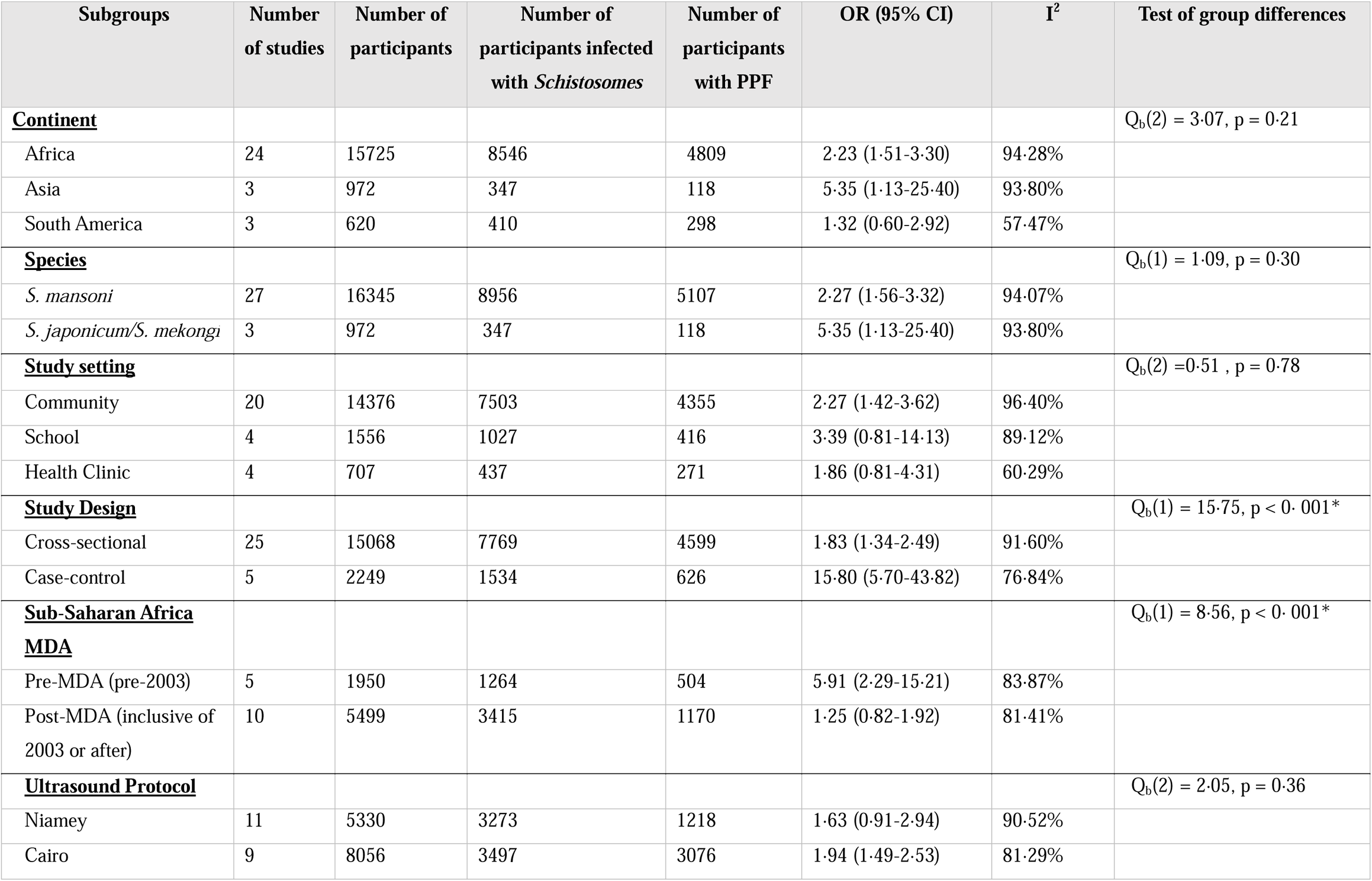
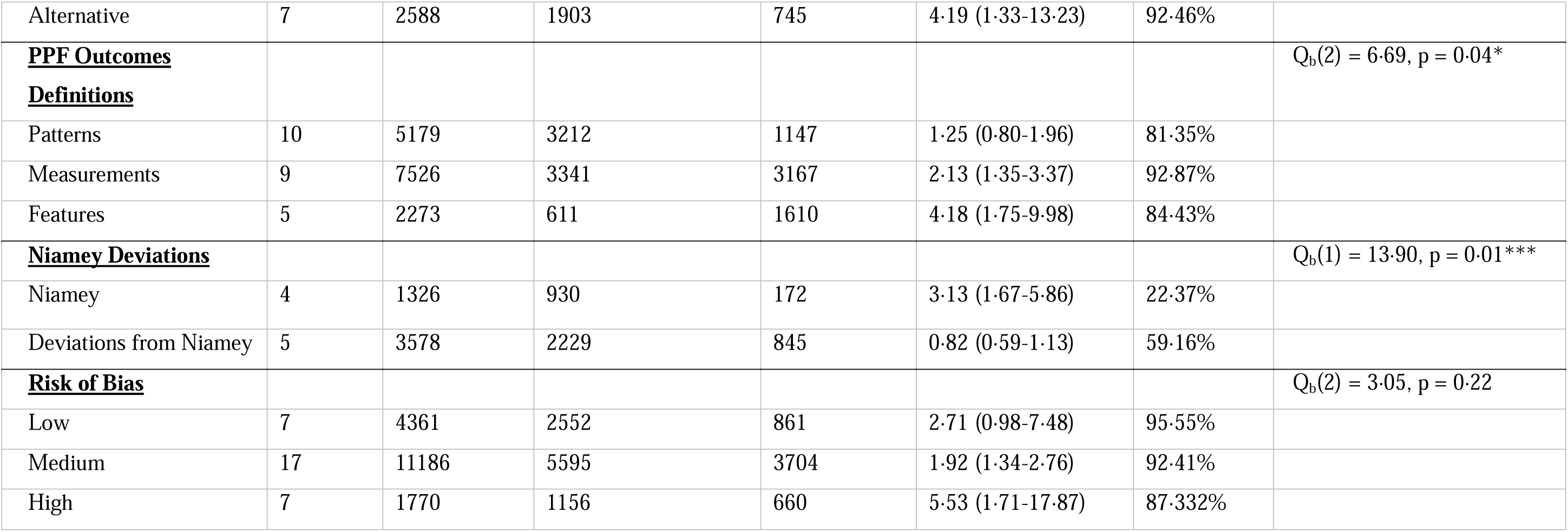
Subgroup analyses.

Protocols for diagnosing PPF by ultrasound varied widely across studies in their use and association with current schistosome infection. No association of current infection status was observed with PPF in studies that used the Niamey protocol (OR 1·63, 95% CI:0·91-2·94). The Cairo protocol (OR 1·94, 95% CI:1·49-2·53) or alternative protocols including Homeida, Managil, Doehring-Schwerdtfeger and Abdel-Wahab (OR 4·19, 95% CI:1·33-13·23) found diagnoses of PPF that were strongly associated with current infection status. Studies were regrouped based on what ultrasound elements were used to define PPF as opposed to the overall protocol as each protocol may contain multiple elements of patterns, measurements, and features. Only measurements (OR 2·13, 95% CI:1·35-3·37) and features (OR 4·18, 95% CI:1·75-9·98) showed a significant association with current infection status. For the Niamey protocol, both patterns and measurements are recommended. Pooled effect sizes were investigated for studies that deviated from the full recommended protocol in any way where authors explicitly stated a modification to the protocol, e.g. categorised PPF grades as B-F instead of C-F or only used patterns. Only studies that strictly followed the full Niamey protocol showed an association of PPF with current infection status, which was of high magnitude with individuals with current schistosome infections over three times more likely to have PPF (95% CI:1·67-5·86).

The high heterogeneity within the pooled analyses (I^2^=94**·**80%) was partially explained by several subgroups (Table 3). Yet, heterogeneity was lowest for subgroups that showed no relationship between current infection status with PPF such as the region of South America (I^2^=57**·**47%), studies in health clinics (I^2^=60**·**29%), and diagnoses that used the Niamey protocol but deviated from the written guidance and procedures (I^2^=22**·**37%).

In total, 26**·**66% (8/30) of studies reported a measure of intensity of schistosome infection against PPF. However, only 16**·**66% (5/30) of studies provided similar measurements of current schistosome infection intensity to enable a meta-analysis. One study only reported the unadjusted values for infection intensity^23^ and another study only reported the adjusted effect size.^52^ Pooled effect sizes are shown in Figure 3a-3b for unadjusted (13·33%, 4/30) and adjusted analyses (13·33%, 4/30), respectively (appendix 1, p28). All analyses showed insignificant relationships between current infection intensity with PPF and no significant differences in pooled effect sizes between infection intensity categories.

**Figure 3:**
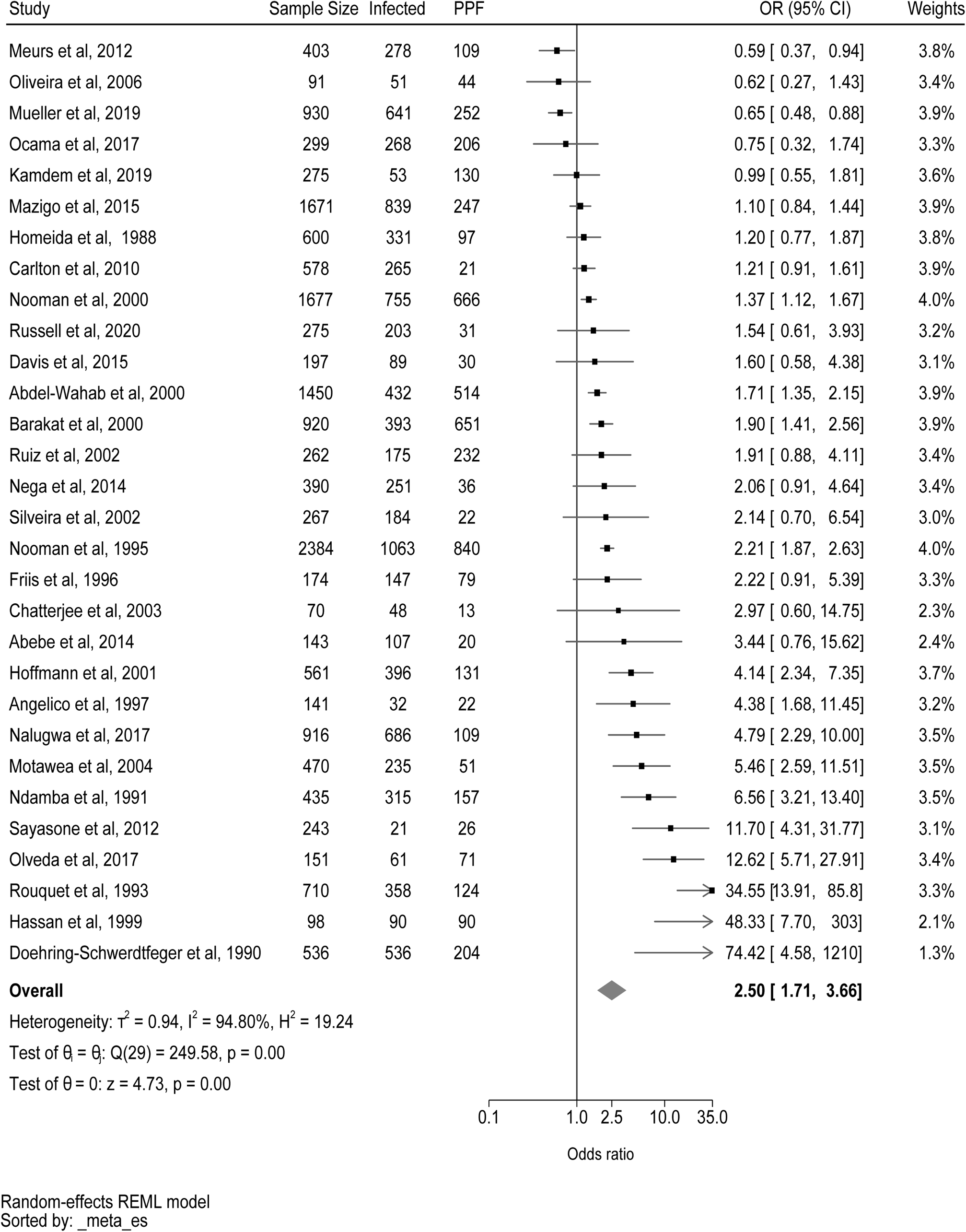
Forest plot of association of current schistosome infection on periportal fibrosis outcome.

**Figure 4:**
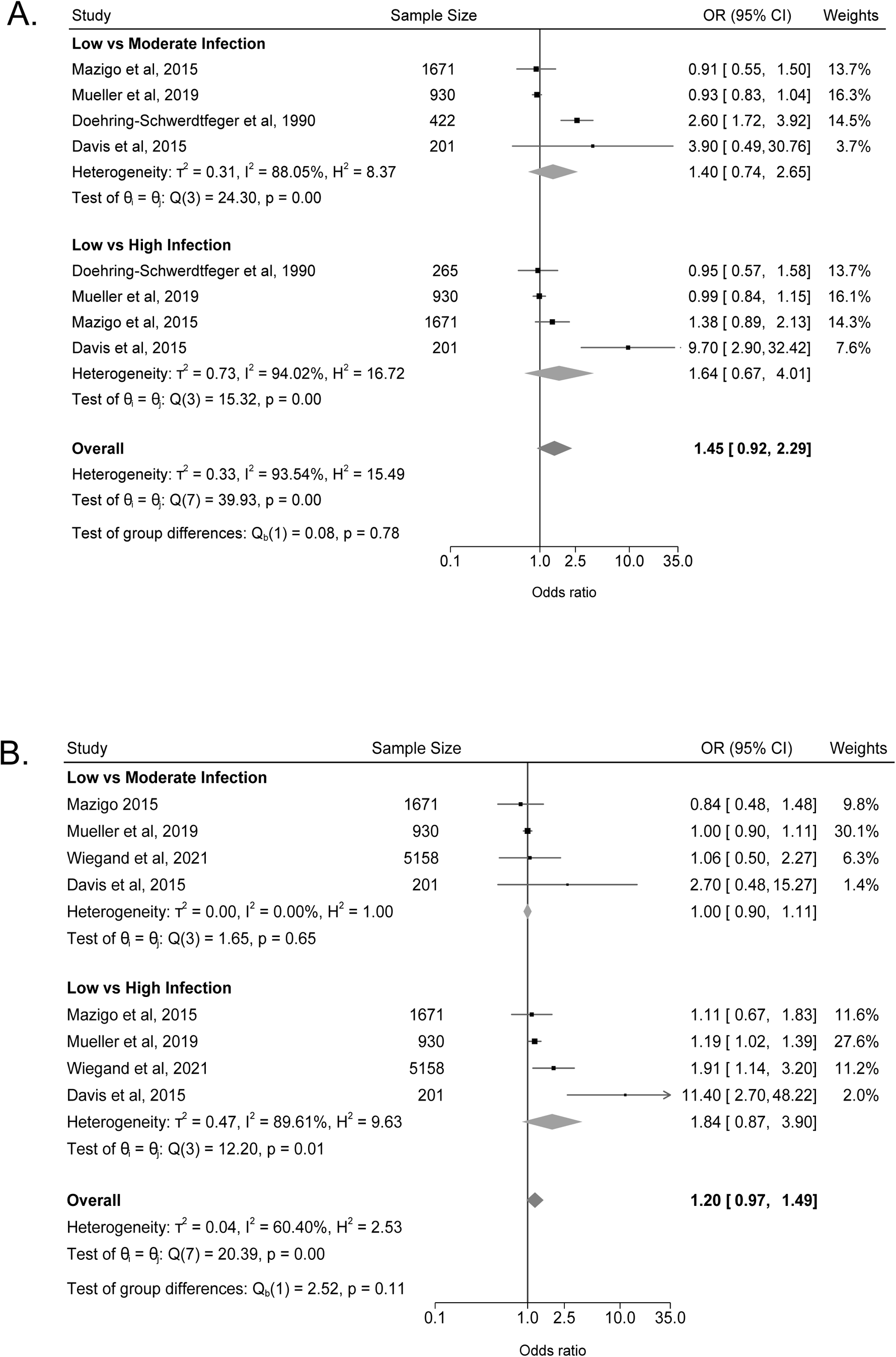
Forest plot of association of current schistosome infection intensity on periportal fibrosis outcome.

Most studies scored a moderate to high ROB (83·7%, 31/30) (appendix 1, pp 29-41). Common reasons for being categorised as moderate or high risk of bias were providing the data in an unadjusted model or only presenting descriptive statistics (93·5%, 29/31) The 30 studies included in the meta-analysis were subject to sensitivity analyses. High (7),^22,24,26,36,46,48,50^ moderate (16),^18,20,21,25,28–30,34,38,40,42–44,51,53,54^ and low (7)^19,23,33,37,39,47,49^ ROB studies were treated as individual subgroups and the pooled effect size of current infection status against PPF was reanalysed (Table 3). The low ROB studies showed no association of current schistosome infection status with PPF (2**·**71, 95% CI:0·98-7·48) with a significant association in the moderate ROB subgroup (OR 1**·**92, 95% CI: 1**·**34-2**·**76) and the strongest association in high ROB subgroup (OR 5**·**53, 95% CI: 1**·**71-17**·**87). No clear indications of publication bias or small study effects were observed (appendix 1, p 42).

## DISCUSSION

WHO guidelines use current infection intensity cut-offs of <5% or <1% prevalence of heavy infection intensities as proxy indicators of schistosomiasis-related morbidity to monitor control and elimination, respectively, as a public health problem.^6^ We synthesised 37 studies and completed a meta-analysis of 30 of those studies that included 17317 participants from 17 countries endemic with *S. mansoni, S. japonicum, or S. mekongi.* There was no evidence for using intensity-based guidelines to approximate PPF likelihood. Current schistosome infection status was shown to influence PPF though the relationship was dependent on the study design deployed and was lost after the widespread introduction of MDA in sub-Saharan Africa.

The irrelevance of infection intensity in our study either conveys important information on disease pathogenesis or simply shows the incompleteness of the existing literature. Current infection intensity may not be the best proxy measure of the history of exposure that reflects chronic infection, re-infection, or current cumulative fluke burden—all of which are important to the long timelines needed for PPF development.^7^ Similarly, current infection intensity does not narrow down risk groups for PPF given the greatest burden is concentrated in school-aged children^55^ and not adults who are more likely to have PPF.^7^ However, only five eligible studies reported the different effect sizes between infection intensity and PPF.^23,24,33,37,52^ Additionally, three of five of these studies defined the comparator group as low infection intensity rather than uninfected participants. Three additional studies could not be included in the meta-analysis as they used unique definitions of infection intensity. To pool effects across studies, the WHO classifications of infection intensity by schistosome species should be reported in future studies.^6^ There is yet insufficient evidence to support WHO guidelines based on using current infection intensity for schistosomiasis-morbidity monitoring with respect to PPF.

Individuals with current schistosome infection were 2·50 times as likely to have PPF compared to currently uninfected individuals. This supports current WHO guidelines for MDA implementation, which prioritise treatment to communities with a high schistosomiasis prevalence, as opposed to guidelines for morbidity monitoring.^6^ This pooled effect size was greater in magnitude than indicators of the history of exposure, such as fishing occupations. Still, it was similar in magnitude to suggested effect sizes for other co-infections or co-morbidities causing liver fibrosis, such as the history of HIV infections or hepatitis B.^7^ Yet, with only a single study^23^ providing adjusted effect sizes eligible for meta-analysis, the pooled effect of 2·50 may be an overestimation of the association of current infection status with PPF. The lack of adjustment also might explain the high heterogeneity observed in the pooled effect for current infection status.

The influence of current infection status on PPF largely was dependent on context and study design. A significant positive association only was observed in African and Asian regions and when participant sampling was directly from communities as opposed to schools or health clinics. The choice of overall study design affected the magnitude of the relationship between current infection status with PPF. Case-control studies have the potential to highlight risk factors for uncommon disease outcomes, such as the latest stages of PPF. Yet, in all five studies reporting a case-control design, none exploited this aspect of case-control design, choosing instead to match on infection exposure, age, and sex as opposed to matching on disease outcomes to retrospectively ascertain the relevance of a wide range of risk factors. ^24,26,36,38,47^ Hence, it is questionable whether the larger effect size in case-control designs represents a true effect size of infection status. Future studies should consider case-control designs that match the disease outcome, especially within health facilities, where patients with pre-diagnosed, severe stages of the disease can be easily identified. Such a monitoring approach may be feasible within existing resource constraints of health systems.

Ultrasound is the primary tool to diagnose PPF with most studies in this meta-analysis and the wider literature, applying the currently WHO-recommended Niamey protocol.^1,4^ The implementation of the Niamey protocol, including the choice of which elements to use and the degree to which the protocol was followed as written affected the association of current infection status with PPF. However, we found no evidence that the Niamey protocol was better at detecting an association of current infection status with PPF as compared to other protocols, such as the Cairo protocol. The significance of the association was lost when examining studies that only used the Niamey protocol. A recent systematic review by Ockenden et al. highlights challenges with existing standardisation of the Niamey protocol including the lack of ultrasound image acquisition guidance, selective subjective usage of specific elements of the protocol, and the lack of quality control and validation.^1^ Infected individuals were over three times more likely to have PPF compared to uninfected individuals when they were diagnosed by studies adhering to the full Niamey protocol. When studies deviated or did not complete the full Niamey protocol the association was removed.

Methods to classify and grade PPF have evolved over the past 40 years and varied based on features, patterns, and measurements.^1^ When these elements were compared across all reported ultrasound protocols, patterns despite the most used element of the Niamey protocol, were uninformative for PPF when compared to measurements (such as portal vein diameter) or features (such as echogenic streaks).^1^ Image patterns, although intuitive, are subject to personal interpretation.^56^ There is a need to review and potentially update the image patterns used for diagnosing schistosomal PPF. Notably, the prevalence of PPF varies based on the protocol and PPF criteria used.^47^ Although measurements showed an association with PPF, the accuracy of measurements depends on the level of clinical expertise of the sonographer. A study by Hoffman et al. showed that for measurements, a high level of clinical expertise is needed to identify the anatomy of the portal vasculature and that sub-segmented and segmented portal branches are commonly confused and, the inclusion of sub-segmental branches could potentially more than triple the percentage of PPF cases.^28^ Improvements in ultrasound technology may help reduce these mistakes. These two examples show that protocols need to be followed and there are clear differences in PPF ascertainment between protocols.

We found that only before the year 2003, i.e. before the widespread introduction of MDA in sub-Saharan Africa,^57^ was a significant association observed between current infection status and PPF. This loss of association between current infection status and PPF after 2003 for studies conducted within sub-Saharan Africa might be attributed to the successful reduction in schistosome infection prevalence after repeated MDA yet with residual morbidities remaining after treatment with praziquantel.^57^ Alternatively, MDA may have introduced heterogeneity into the known infection epidemiology in endemic populations due to systematically missing individuals across multiple rounds of MDA.^10^

A limitation of our review was that we excluded non-English language studies at the full test screening stage; 46 non-English reports were excluded with the majority of these published in Mandarin. Therefore, only three *S. japonicum* and *S. mekongi* reports were included. This likely underrepresents these species, studies conducted in Asia, and studies that use the China Centres for Disease Control protocol for *S. japonicum* diagnoses.^1^ As we did not limit our search string to periportal fibrosis, it was unlikely that interseptal fibrosis was missed in English studies.

There were many limitations of the existing literature. Some studies used parenchymal fibrosis and periportal fibrosis interchangeably introducing heterogeneity into the PPF definitions.^53,58^ PPF likelihood changes across the lifespan of an individual; increasing in adolescents, remaining stable in adulthood, and declining above 55 years of age.^7^ However, many of the included studies either did not report age ranges or only reported one wide age range such as 1-95 years.^37^ Exposure and PPF outcomes were not broken down by age, so no sub-group analyses by age were possible and remain an area for future study. Most studies were of high or moderate risk of bias with adjusted analyses considering common confounders of age and sex rarely reported and insufficiently shared to calculate any adjusted pooled effect sizes. More studies with adequate study designs to address the association of current schistosome infection with PPF are required. Current infection status with intestinal schistosomes was tenuously associated with PPF, but no association was observed for current infection intensity. Further epidemiological studies, that adjust for age and gender, which capture the influence of current, chronic, and historical schistosome infection on PPF are required. At present, there is no support for approximating PPF using current schistosome infection intensity, and infection status appears irrelevant within the context of repeated MDA that is now routine in areas endemic with schistosomiasis. For these reasons, there is an urgent need to directly monitor PPF-related morbidity in areas endemic to schistosomiasis and to develop strategies to enable such monitoring within already constrained health systems.

## Data Availability

The study is a systematic review and meta-analysis of publicly available literature.

## Acknowledgements

We thank the SchistoTrack Research Group for wider support on meta-analyses and feedback during group meetings.

## Author contributions

Resources and supervision: GFC. Conceptualization: GFC. Data curation: AE, LW, DT. Pre-registration protocol, data extraction tool, and risk of bias tool: AE, RM, GFC. Data validation: LW, NR, RM. Investigation and methodology: AE, LW, RM, GFC. Formal analysis and visualization: AE, LW. Writing – original draft: AE, LW, GFC. Writing – review and editing: AE, LW, DT, NR, DT, GFC. Funding acquisition: GFC.

## Open access statement

This research was funded in whole, or in part, by the UKRI EPSRC [EP/X021793/1]. For the purpose of Open Access, the author has applied a CC BY public copyright licence to any Author Accepted Manuscript version arising from this submission.

## Appendices

Appendix 1: Supplementary material

Appendix 2: Data extraction table

