## Appendix 1 for "Association of current *Schistosoma mansoni, S. japonicum,* and *S. mekongi* infection status and intensity with periportal fibrosis: a systematic review and meta-analysis"

**Title**

Text S1: search strategy

We searched 5 databases on 18th May 2022, Cochrane Central Register of Controlled Trials (Cochrane Library, Wiley)[Issue 4 of 12, April 2022), Embase (OvidSP)[1974-present], Global Health (OvidSP)[1973 to 2022 Week 19], Global Index Medicus (https://pesquisa.bvsalud.org/gim/ ) and Medline (OvidSP)[1946-present]. An updated search was conducted on 24th August 2022. We developed a search strategy combining free-text keywords and subject headings for our key concepts of schistosomiasis and periportal fibrosis. We applied study filters for controlled studies developed by the Cochrane Effective Practice of Care Group and observational studies developed by Scottish Intercollegiate Guidelines Network. No date or language limits were applied, but we excluded animal studies where possible. References were exported to Endnote where a further search for animal studies was conducted so that they could be excluded. The remaining studies were exported to Covidence for screening.

Table S1: Summary of number search hits across five databases

| **Database** | **Interface** | **Coverage** | **Hits: 18/05/22** | **Hits: 24/08/22** | **Total** |
| --- | --- | --- | --- | --- | --- |
| Cochrane Central Register of Controlled Trials | Cochrane Library, Wiley | Issue 4 of 12, April 2022 | 20 | 11 | 31 |
| Embase | OvidSP | 1974-present | 526 | 159 | 685 |
| Global Health | OvidSP | 1973 to 2022 Week 19 | 542 | 261 | 803 |
| Global Index Medicus | <https://pesquisa.bvsalud.org/gim/> |  | 339 | 359 | 698 |
| Medline | OvidSP | 1946-present | 450 | 247 | 697 |
| **Total** |  |  | **1877** | **1037** | **2914** |
| Duplicates - removed in Covidence |  |  |  |  | 902 |
| Animal studies excluded in Endnote |  |  |  |  | 286 |
| **Final total** |  |  |  |  | **1726** |

Table S2: Summary of search terms for Medline

| [Medline (Ovid MEDLINE® Epub Ahead of Print, In-Process & Other Non-Indexed Citations, Ovid MEDLINE® Daily and Ovid MEDLINE®) 1946 to present](https://ovidsp.ovid.com/ovidweb.cgi?T=JS&NEWS=N&PAGE=main&SHAREDSEARCHID=hIDMlca3BhMVc5e3huENHd4JGbJXDMGypJ8WizTMeVyGPJEiQbKd7V93eUorNSpb) | 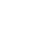<https://ezproxy-prd.bodleian.ox.ac.uk:2483/ovid-a/ovidweb.cgi?&S=DGLOFPGFPGEBIFGIJPPJBHBFJPFIAA00&R=11&Search+Annotations+Options=SA> |
| --- | --- |
| schistosomiasis/ or schistosomiasis japonica/ or schistosomiasis mansoni/ | 22799 |
| schistosoma/ or schistosoma japonicum/ or schistosoma mansoni/ | 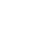<https://ezproxy-prd.bodleian.ox.ac.uk:2483/ovid-a/ovidweb.cgi?&S=DGLOFPGFPGEBIFGIJPPJBHBFJPFIAA00&R=13&Search+Annotations+Options=SA>16729 |
| (schistosomiasis or bilharzia or snail* fever).ti,ab,kf. | 19616 |
| (schistosoma* adj2 (mansoni or japonic* or mekongi)).ti,ab,kf. | 15730 |
| (s mansoni or s japonic* or s mekongi).ti,ab,kf. | 7945 |
| 1 or 2 or 3 or 4 or 5 | 35064 |
| Liver Cirrhosis/ | 81896 |
| Fibrosis/ | 37787 |
| ((periportal or peri-portal or portal or liver or hepati*) adj2 fibrosis).ti,ab,kf. | 30109 |
| ((periportal or peri-portal or portal or liver or hepati*) adj2 cirrho*).ti,ab,kf. | 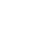<https://ezproxy-prd.bodleian.ox.ac.uk:2483/ovid-a/ovidweb.cgi?&S=DGLOFPGFPGEBIFGIJPPJBHBFJPFIAA00&R=29&Search+Annotations+Options=SA>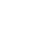<https://ezproxy-prd.bodleian.ox.ac.uk:2483/ovid-a/ovidweb.cgi?&S=DGLOFPGFPGEBIFGIJPPJBHBFJPFIAA00&R=31&Search+Annotations+Options=SA>52809 |
| symmer*.ti,ab,kf. | 351 |
| 7 or 8 or 9 or 10 or 11 | 154442 |
| 6 and 12 | 1775 |
| randomized controlled trial.pt. | 575687 |
| controlled clinical trial.pt. | 95003 |
| pragmatic clinical trial.pt. | 2141 |
| multicenter study.pt. | 324960 |
| non-randomized controlled trials as topic/ | 1048 |
| interrupted time series analysis/ | 1689 |
| controlled before-after studies/ | 704 |
| (randomis* or randomiz* or randomly).ab. | 1026092 |
| groups.ab. | 2398214 |
| (trial or multicenter or multi center or multicentre or multi centre).ti. | 323846 |
| (intervention? or effect? or impact? or controlled or control group? or (before adj5 after) or (pre adj5 post) or ((pretest or pre test) and (posttest or post test)) or quasiexperiment* or quasi experiment* or evaluat* or time series or time point? or repeated measur*).ti,ab. | 11189443 |
| or/14-24 | 12430283 |
| exp animals/ not humans.sh. | 5040164 |
| 25 not 26 | 10158189 |
| 13 and 27 | 603 |
| Epidemiologic studies/ | 9159 |
| exp case control studies/ | 1348264 |
| exp cohort studies/ | 2387291 |
| **Case control.tw.** | 145905 |
| (cohort adj (study or studies)).tw. | 282899 |
| Cohort analy$.tw. | 10658 |
| (Follow up adj (study or studies)).tw. | 54255 |
| (longitudinal or retrospective or prospective).tw. | 1557400 |
| cross sectional.tw. | 464441 |
| Cross-sectional studies/ | 437793 |
| 29 or 30 or 31 or 32 or 33 or 34 or 35 or 36 or 37 or 38 | 3701156 |
| exp animals/ not humans.sh. | 5040164 |
| 39 not 40 | 3621636 |
| 13 and 41 | 236 |
| 28 or 42 | 692 |

Table S3: Summary of search terms for Embase

| [Embase 1974 to present](https://ovidsp.ovid.com/ovidweb.cgi?T=JS&NEWS=N&PAGE=main&SHAREDSEARCHID=2rMjLp7RuSMokpkVYrRoUAvCZDALnhgzuqpJhzim7JUl7bAOz9WS3nilJMqhJRznU) |  |
| --- | --- |
| schistosomiasis/ or schistosomiasis japonica/ or schistosomiasis mansoni/ | 22066 |
| schistosoma/ or schistosoma japonicum/ or schistosoma mansoni/ | 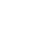<https://ezproxy-prd.bodleian.ox.ac.uk:2483/ovid-a/ovidweb.cgi?&S=DGLOFPGFPGEBIFGIJPPJBHBFJPFIAA00&R=13&Search+Annotations+Options=SA>21040 |
| (schistosomiasis or bilharzia or snail* fever).ti,ab,kf. | 19285 |
| (schistosoma* adj2 (mansoni or japonic* or mekongi)).ti,ab,kf. | 16034 |
| (s mansoni or s japonic* or s mekongi).ti,ab,kf. | 8748 |
| 1 or 2 or 3 or 4 or 5 | 37065 |
| liver fibrosis/ | 55735 |
| ((periportal or peri-portal or portal or liver or hepati*) adj2 fibrosis).ti,ab,kf. | 49737 |
| ((periportal or peri-portal or portal or liver or hepati*) adj2 cirrho*).ti,ab,kf. | 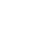<https://ezproxy-prd.bodleian.ox.ac.uk:2483/ovid-a/ovidweb.cgi?&S=DGLOFPGFPGEBIFGIJPPJBHBFJPFIAA00&R=27&Search+Annotations+Options=SA>72359 |
| symmer*.ti,ab,kf. | 126 |
| 7 or 8 or 9 or 10 | 133050 |
| 6 and 11 | 1962 |
| exp randomized controlled trial/ | 724862 |
| controlled clinical trial/ | 466805 |
| multicenter study/ | 333477 |
| single blind procedure/ | 47238 |
| double blind procedure/ | 197819 |
| crossover procedure/ | 71227 |
| time series analysis/ | 33550 |
| pretest posttest control group design/ | 604 |
| (randomis* or randomiz* or randomly).ab. | 1441957 |
| factorial.ti,ab. | 43772 |
| (crossover* or cross over*).ti,ab. | 119442 |
| ((doubl* or singl*) adj blind*).ti,ab. | 260165 |
| (assign* or allocat* or volunteer* or placebo*).ti,ab. | 1187597 |
| (trial or multicenter or multi center or multicentre or multi centre).ti. | 452153 |
| (intervention? or effect? or impact? or controlled or control group? or (before adj5 after) or (pre adj5 post) or ((pretest or pre test) and (posttest or post test)) or quasiexperiment* or quasi experiment* or evaluat* or time series or time point? or repeated measur*).ti,ab. | 14302670 |
| or/13-27 | 15150076 |
| exp animal/ not human/ | 5130795 |
| 28 not 29 | 12634751 |
| 12 and 30 | 610 |
| Clinical study/ | 160060 |
| **case control study/** | 191770 |
| family study/ | 25675 |
| longitudinal study/ | 176956 |
| retrospective study/ | 1293209 |
| prospective study/ | 788075 |
| cohort analysis/ | 882925 |
| ((Cohort or case control or follow up or observational or epidemiologic or cross sectional) adj (study or studies)).mp. | 1560572 |
| or/32-39 | 3687606 |
| exp animal/ not human/ | 5130795 |
| 40 not 41 | 3638847 |
| 12 and 42 | 202 |
| 31 or 43 | 679 |

Table S4: Summary of search terms for Cochrane Central register for Controlled Trials

Cochrane Central Register of Controlled Trials

MeSH descriptor: [Schistosomiasis] this term only

MeSH descriptor: [Schistosomiasis japonica] explode all trees

MeSH descriptor: [Schistosomiasis mansoni] explode all trees

MeSH descriptor: [Schistosoma] this term only

MeSH descriptor: [Schistosoma japonicum] explode all trees

MeSH descriptor: [Schistosoma mansoni] explode all trees

((schistosomiasis or bilharzia or "snail fever")):ti,ab,kw OR ((schistosoma* NEAR/2 (mansoni or japonic* or mekongi))):ti,ab,kw OR (("s mansoni" or "s mekongi" or "s japonica" or "s japonicum")):ti,ab,kw

#1 or #2 or #3 or #4 or #5 or #6 or #7

MeSH descriptor: [Liver Cirrhosis] this term only

MeSH descriptor: [Fibrosis] this term only

(((periportal or peri-portal or portal or liver or hepati*) NEAR/2 fibrosis)):ti,ab,kw OR (((periportal or peri-portal or portal or liver or hepati*) NEAR/2 cirrho*)):ti,ab,kw OR (symmer*):ti,ab,kw

#9 or #10 or #11

#8 and #12

Table S5: Summary of search terms for Global Index Medicus

| Global Index Medicus |  |
| --- | --- |
| Title, abstract, subject = ((schistosoma OR schistosomiasis OR bilharzia OR "snail fever") AND (liver OR hepat* OR periportal OR peri-portal OR portal) AND (cirrhosis OR cirrhotic OR fibrosis OR fibrotic OR symmer*)) | 698 |

| P | Population | - Participants could be of any age or sex - Participants could be individuals living in regions where *S. mansoni, S. japonicum,* or *S. mekongi* are endemic |
| --- | --- | --- |
| E | Exposure | Current infection status and current intensity of infection   - Current infection status (binary variable, yes/no) is determined by the presence of schistosome eggs (at least one egg per gram of stool), or a positive result from an antibody-based or antigen-based diagnostic test. - Infection intensity can be defined by the total number of eggs per gram (EPG) of stool, which can be classified into WHO categories of not infected, low intensity, moderate intensity, and heavy intensity. If WHO categories were not provided, but infection intensity was presented in the form of continuous egg counts then this information was still collected. |
| C | Comparator | - Uninfected individuals were the comparator for infection status - Individuals of lower-category infection intensities were the comparators for moderate and high-intensity infection. |
| O | Outcome | The outcome is periportal or Symmer’s pipestem fibrosis as defined by the study authors and as determined by ultrasound examination (binary, yes/no)   - If liver image pattern information was provided, this was used as a binary/categorical indicator of the direct grading of the liver pattern. |

Table S6: PECO inclusion criteria

| **Variable** | **Definition/Units** |
| --- | --- |
| Study ID | Author and year of publication |
| Author | First author |
| Year of publication |  |
| Journal | Journal of publication |
| Study design | Cohort/Case-control/Randomised control trial/cross sectional |
| Study aim | Main aim of the study |
| Study year | Year that the study was performed if included in the text. If not, ‘Not reported’ |
| Study duration | How long the study was ran for. |
| Country | List all the countries if multiple |
| Study locality | urban, peri-urban, urban. If not stated, implied by reviewers |
| Study setting | community, healthcare setting, school, workplace |
| Total sample size | Number of participants included in the study. |
| Participant age | As reported |
| Participant sex | As reported. If not stated both male and female implied |
| Total number of individuals with intestinal *Schistosoma* infection | Currently diagnosed |
| Total number of individuals with periportal fibrosis | As diagnosed by ultrasound |
| Treatment status (with praziquantel) |  |
| *Schistosoma* species | *S. mansoni, S. japonicum, S. mekongi* |
| Diagnostic tool (exact) | As stated |
| Infectious status definition | As stated |
| Infection intensity definition | As stated |
| Total individuals in each WHO infection intensity category |  |
| Author defined Periportal fibrosis definition | As stated |
| Ultrasound Protocol | Cited or stated in text |
| Unadjusted and adjusted odds ratios, relative risks, hazard ratios and corresponding 95% confidence intervals or the necessary information to calculate the effect size |  |
| Any relevant covariates |  |

Table S7: Variable extracted from the studies, with definitions

Text S2 Subgroup Variables

Continent based upon the study country for each study, based on world bank definitions of continent. These were categorically coded as Africa, Asia, and South America.

Species were categorically codes as *S. mansoni, S. japonicum,* or, *S. mekongi* extracted based on the study author description.

Study setting was either explicitly stated or inferred by the reviewers. Categorised into urban, peri-urban or rural.

Before/after MDA, these restricted studies only in performed in Sub-Saharan Africa (defined by world bank). These were coded as binary before/after. Studies from north Africa e.g. Egypt and Sudan were excluded in this subgroup. MDA was introduced in sub-Saharan Africa in 2003 (Deol *et al*., 2019).^55^ If study year was not explicityly states, reviewers inferred the study year based upon the publication date i.e. 2014 publication date was categories as after 2003. In the case of uncertainly e.g. 2004 publication year, this was coded as uncertain.

Ultrasound protocol was extracted either based on authors explicitly stating or describing the name of the protocol or referencing/citing the protocol in their methods. If they described a protocol but did not name it, these were classified as undefined. If they had no mention or description of an ultrasound protocol these were classified as not stated. If authors stated they used one type of protocol but described a protocol that did not align with this name, the authors name of the protocol was extracted as the ultrasound definitions were also extracted.

Ultrasound outcome definitions were based upon definitions by Ockenden *et al.,* 2024.^1^Although ultrasound protocols are associated with different ultrasound elements, protocols can use different elements to grade fibrosis e.g. Niamey combines patterns and measurements to generate a score. Measurements: measurements of liver anatomy such as vessel diameters or organ size. Feature: consider distinct characteristics such as liver surface texture or echogenic streaks Patterns: using the overall pattern of a liver e.g. starry sky, birds claw. Typically aligns with the Niamey protocol

Niamey deviations were restricted to only Niamey protocols and only Niamey protocols that provided some description of the methods they used. For example, studies that only cited the protocol were not included. Followed the protocol meant that both patterns and measurements were taken and a score was generated as according to the original protocol. Deviations for example, re classifying fibrosis as B-F rather than C-F or only using image patterns and not measurements or explicitly stating modified Niamey protocol.

Table S8: Risk of Bias tool adapted from National Institutes of Health Tool from the National Heart, Lung, and Blood Institute for observational cohort and cross-sectional studies.

| **Category** | **Questions** | **Guidance on when to select 'yes'** | **Guidance on when to select 'no'** |
| --- | --- | --- | --- |
| Aim | D1 Was the research question or objective in this paper clearly stated? | Authors clearly describe the goal of their research, e.g. the aim was to estimate the association between *Schistosoma mansoni, S. japonicum and S. mekongi* infection status and/or intensity and liver fibrosis by ultrasound (USS) | No aim, unspecific aim, aim does not match the analyses completed or aim does not match the outcomes reported |
| Representativeness | D2 Was the study population clearly specified and defined? | Authors describe the group of people from which the study participants were selected or recruited, using demographics, location, and time period, e.g. the study population were male and female individuals of any age living in regions where *S. mansoni, S. japonicum, or S. mekongi* are endemic. | Description lacking specifics on study population demographics, location, and time period |
|  | D3 Was the method for selecting the sample clearly described? | Sampling method clearly described, e.g. 10 school-age children (5-14 years old) per class in each of the five schools were sampled using stratified random sampling | Lacking details to understand how sampling was done, e.g. 50 adults per village were selected for inclusion in the study |
|  | D4 Response rate >50% or differences between respondents and non-respondents described? | Either 1) Provides statistics on response rate, e.g. 90% of responders selected from village registries agreed to participate in the study, or 2) Provides differences in known characteristics of responders (age, gender) ideally assessed with the appropriate statistical test | Low response rate, lacking information on response rate and/or differences between responders and non- responders provided |
|  | D5 Is the sample representative of the population from which it is drawn (including similar timeframe)? Were inclusion and exclusion criteria for being in the study pre-specified and applied uniformly to all participants? (For case-control studies: Were the cases clearly defined and differentiated from controls?) | Clear inclusion and exclusion criteria. Selected participants representative of the population they are sampled from (and in cohort or case-control studies, both groups selected from same underlying population), e.g. random sample from community    (For case-control studies: cases and controls are clearly defined, cases and controls are drawn from the same population and there are no significant differences between the cases and controls) | Sample not representative of population, e.g. children sampled in schools may not be representative of school-age children, or participants selected based on liver fibrosis may not be representative of village population (e.g. higher social status households, all households with home latrines, etc.). Excluded participants without providing a reason.    (For case-control studies: no clear definitions of cases and/or controls, cases and controls are not matched or stratified, despite being specified in the design) |
| **Category** | **Questions** | **Guidance on when to select 'yes'** | **Guidance on when to select 'no'** |
|  | D6 Was a sample size justification, power description, or variance and effect estimates provided? | Authors present their reasons for selecting or recruiting the number of people included or analysed, e.g. state that they performed power calculations before selecting sample | No justification on size of selected sample provided |
| Exposure | D7  Were the exposures of interest measured prior to the outcome being measured?  Was the timeframe sufficient so that one could reasonably expect to see an association between exposure and outcome if it existed? | Information relating to liver fibrosis was obtained post infection, i.e. the investigator can confirm that the exposure occurred prior to the development of periportal fibrosis | The investigator cannot confirm that the infection occurred prior to the development of periportal fibrosis, e.g. authors measured infection status after the detection of liver fibrosis by ultrasound. |
|  | D8 Did the study define the exposures? Did the study examine different levels (i.e. more than two levels) of infection intensity as related to the outcome (liver fibrosis)? | The study describes infection status clearly as having an eggs per gram of stool (EPG) count of >0 EPG with a form of quality control measure, e.g. ensuring a senior laboratory technician re-analyses 10% of stool sample using the Kato-Katz microscopy technique or analyses multiple urine samples. For the use of a commercial diagnostic for antigen-based and antibody-based tests, analyses should be done in a reference laboratory.    Study describes variable construction and clearly defined levels of infection intensity and defines unit of measurement, e.g.  1-99 EPG, 100-399 EPG, >400 EPG | Study does not explicitly define infection status, constructs a binary variable of infection intensity or doesn't define variable construction and measurement unit, e.g. infection intensity/no intensity |
|  | D9 Were the assessors of exposure blinded to the case or control status of participants? | The laboratory technicians were not aware of the periportal fibrosis stratus of the participants and/or the USS trained personnel were not aware of the infection status | The laboratory technicians were aware of the periportal fibrosis status of the participants and/or the USS trained personnel were aware of the infection status |
| Outcomes | D10 Was the outcome (dependent variable) clearly defined, valid, reliable, and implemented consistently across all study participants, i.e. measured by trained personnel? | Clearly states that at least two USS measurements were taken by a trained sonographer, using validated methodologies from either the Niamey or Cairo. | The study takes fewer than 2 USS measurements taken, a specified protocol is not followed for the liver ultrasonography, or the individuals taking the USS measurements are untrained or other allied healthcare professionals that are not trained |
| **Category** | **Questions** | **Guidance on when to select 'yes'** | **Guidance on when to select 'no'** |
|  | D11 Were exposures and outcomes not measured by the same person, i.e. were the outcome assessors blinded to the exposure status of participants? | Infection status and/or intensity and USS done by different people, e.g. infection status using microscopy (exposure) and USS by a trained sonographer (outcome) | Same person measuring infection status and liver fibrosis, e.g. sonographers (outcome measurement) recruited to measure infection status (exposure) |
|  | D12 Was loss to follow-up after baseline 20% or less? Is there missing data? | Follow-up in studies with multiple rounds of data collection not less than 80%, e.g. attrition at follow-up was 5% | Loss to follow-up was 20% or more. Or if there were 200 participants in the study but only 100 in the model, missingness is >20% and therefore unacceptable |
| Covariates | D13 Were the covariates clearly defined, valid, reliable, and implemented consistently across all study participants? | For example age categories 5-14 years, 15-18 years, 18+ years | Covariates lacking information on definitions, e.g. PSACs, SACs, adults (and no definition of respective age range) |
| Analysis | D14 Were adjusted (inclusive of at least age and gender) and unadjusted effect measures and their 95% CIs reported? | Studies report both types of models and report on the covariates adjusted for, e.g. unadjusted association between high infection intensity and liver fibrosis status when compared to low infection intensity was OR = 2.20, 95% CI 2.12, 2.34, when adjusting for gender the association was OR = 1.97, 95% CI 1.94, 2.06 | Studies report only adjusted or unadjusted ORs/RRs, fail to provide 95% CIs or descriptions of variables adjusted for, e.g. OR for liver fibrosis for respondents with high intensity infection status compared to low intensity infection status was OR = 2.20. |

| Table S9: Excluded Full Text reports. |  |  |
| --- | --- | --- |
| Title | Published Year | Notes |
| Effectiveness of FibroTouch combined with four hepatic fibrosis biomarkers for evaluation of the liver fibrosis degree among patients with chronic schistosomiasis-induced liver disorders | 2022 | Exclusion reason: Study not in English; |
| Schistosoma mansoni infection and the occurrence, characteristics, and survival of patients with hepatocellular carcinoma: an observational study over a decade | 2022 | Exclusion reason: Wrong exposure; |
| The Coutinho index as a simple tool for screening patients with advanced forms of Schistosomiasis mansoni: A validation study | 2022 | Exclusion reason: No comparator; |
| Cross-sectional survey between schistosomiasis liver fibrosis and health-related quality of life among agricultural workers | 2021 | Exclusion reason: Study not in English; |
| Phenotypic Characterization of CD4+ T Lymphocytes in Periportal Fibrosis Secondary to Schistosomiasis | 2021 | Exclusion reason: Not enough information for outcome effect sizes; |
| Evaluation of Schistosomiasis Mansoni Morbidity by Hepatic and Splenic Elastography | 2021 | Exclusion reason: No comparator; |
| Regression of Schistosoma mansoni associated morbidity among Ugandan preschool children following praziquantel treatment: A randomised trial | 2021 | Exclusion reason: No comparator; |
| Superiority of rectal snip over serology in detection of schistosomiasis eradication: A pilot study | 2021 | Exclusion reason: Outcome not binary; |
| The dynamics of hepatic fibrosis related to schistosomiasis and its risk factors in a cohort of China | 2021 | Exclusion reason: Not enough information for outcome effect sizes; |
| Changing trends of schistosome infection and liver fibrosis among residents in the Poyang Lake region | 2021 | Exclusion reason: Study not in English; |
| Schistosoma mansoni-related periportal fibrosis; can we use APRI and PSDR levels in the real-time selection of patients for targeted endoscopy in a resource-limited setting? A case-control study | 2021 | Exclusion reason: Not enough information for outcome effect sizes; |
| Association of IL-9, IL-10, and IL-17 Cytokines with Hepatic Fibrosis in Human Schistosoma mansoni Infection | 2021 | Exclusion reason: No comparator; |
| The Coutinho index as a simple tool for screening patients with advanced forms of Schistosomiasis mansoni: a validation study | 2021 | Exclusion reason: Duplicate; |
| Persistent Colonic Schistosomiasis among Symptomatic Rural Inhabitants in the Egyptian Nile Delta | 2021 | Exclusion reason: Not enough information for outcome effect sizes; |
| Serum vitamin d expression in advanced schistosomiasis patients with hepatic fibrosis and its association with disease progression | 2020 | Exclusion reason: Not enough information for outcome effect sizes; |
| Use of a tablet-based system with portable transducers to perform abdominal ultrasounds in a field investigation of schistosomiasis-related morbidity | 2020 | Exclusion reason: Duplicate; |
| Contribution of ultrasonography in the diagnosis of periportal fibrosis caused by schistosomiasis | 2020 | Exclusion reason: Not enough information for outcome effect sizes; |
| Ultrasound evaluation of schistosomiasis-related morbidity among the Xakriaba people in the state of Minas Gerais, Brazil | 2020 | Exclusion reason: Not enough information for outcome effect sizes; |
| Prevalence, exposures, and risk for schistosomiasis in the U.S. military | 2020 | Exclusion reason: Cannot source full text; authors did not respond; |
| Evaluation of hepatic fibrosis by elastography in patients with schistosomiasis mansoni | 2020 | Exclusion reason: No comparator; |
| Can early diagnosis of varices, regular praziquantel, and reduction of hepatitis coinfection reduce mortality among patients attended for periportal fibrosis in northwestern Tanzania? a case-control study | 2020 | Exclusion reason: Wrong outcomes; |
| The Prevalence, Predictors, and In-Hospital Mortality of Hepatic Encephalopathy in Patients with Liver Cirrhosis Admitted at St. Dominic Hospital in Akwatia, Ghana | 2020 | Exclusion reason: Wrong outcomes; |
| Patients with severe schistosomiasis mekongi morbidity demonstrating ongoing transmission in Southern Lao People's Democratic Republic | 2020 | Exclusion reason: No comparator; |
| Ultrasonographic evidence of hepatic disease due to schistosoma mansoni in children in the marolambo district, Madagascar | 2018 | Exclusion reason: Duplicate; |
| Plasma levels of innate immune mediators are associated with liver fibrosis in low parasite burden Schistosoma mansoni-infected individuals | 2018 | Exclusion reason: No comparator; |
| Correlation among three non-invasive methods (APRI, FIB-4 and Transient Elastography) to evaluate liver function and stiffness in patients with viral hepatitis C or schistosomiasis mansoni | 2018 | Exclusion reason: Wrong outcome measurement; |
| Bilharziosis in migration medicine-an underestimated problem? | 2018 | Exclusion reason: Not enough information for outcome effect sizes; |
| Transient elastography evaluation of hepatic and spleen stiffness in patients with hepatosplenic schistosomiasis | 2017 | Exclusion reason: Wrong exposure diagnosis method; |
| Influence of a TNF-alpha Polymorphism on the Severity of Schistosomiasis Periportal Fibrosis in the Northeast of Brazil | 2017 | Exclusion reason: No comparator; |
| Impact of old Schistosomiasis infection on the use of transient elastography (Fibroscan) for staging of fibrosis in chronic HCV patients | 2017 | Exclusion reason: Wrong exposure diagnosis method; |
| Biennial versus annual treatment for schistosomiasis and its impact on liver morbidity | 2017 | Exclusion reason: Not enough information for outcome effect sizes; |
| Schistosoma mansoni infection and its related morbidity among adults living in selected villages of Mara region, north-western Tanzania: a cross-sectional exploratory study | 2017 | Exclusion reason: Not enough information for outcome effect sizes; |
| Co-infection of Schistosoma mansoni/Hepatitis C virus and their associated factors among adult individuals living in fishing villages, north-western Tanzania | 2017 | Exclusion reason: Not enough information for outcome effect sizes; |
| Diagnosis of coinfection by schistosomiasis and viral hepatitis B or C using 1H NMR-based metabolomics | 2017 | Exclusion reason: Wrong exposure diagnosis method; |
| Schistosomiasis and hepatopulmonary syndrome: the role of concomitant liver cirrhosis | 2017 | Exclusion reason: No comparator; |
| Pulmonary shunts in severe hepatosplenic schistosomiasis: Diagnosis by contrast echocardiography and their relationship with abdominal ultrasound findings | 2017 | Exclusion reason: No comparator; |
| New index for the diagnosis of liver fibrosis in Schistosomiasis mansoni | 2017 | Exclusion reason: No comparator; |
| Assessment of the effect of treatment and assistance program on advanced patients with schistosomiasis japonica in China from 2009 to 2014 | 2016 | Exclusion reason: Not enough information for outcome effect sizes; |
| Egy-score can predict portal hypertension in chronic hepatitis C with good accuracy | 2016 | Exclusion reason: Duplicate; |
| Increased Hepatic Arterial Blood Flow Measured by Hepatic Perfusion Index in Hepatosplenic Schistosomiasis: New Concepts for an Old Disease | 2016 | Exclusion reason: No comparator; |
| Morbidity of mansoni schistosomiasis in Pernambuco-Brazil: Analysis on the temporal evolution of deaths, hospital admissions and severe clinical forms (1999-2014) | 2016 | Exclusion reason: Wrong exposure diagnosis method; |
| Is ultrasonography useful for population studies on schistosomiasis mansoni? An evaluation based on a survey on a population from Kome Island, Tanzania | 2016 | Exclusion reason: Outcome not binary; |
| [Expression of Tim-3 on Peripheral CD56(+) NK Cells and Its Correlation with Liver Fibrosis in Patients with Advanced Schistosomiasis] | 2015 | Exclusion reason: Study not in English; |
| Transient elastography evaluation of hepatic and spleen stiffness in patients with hepatosplenic schistosomiasis | 2015 | Exclusion reason: Duplicate; |
| Can mass drug administration lead to the sustainable control of schistosomiasis? | 2015 | Exclusion reason: Not enough information for outcome effect sizes; |
| Impact of old schistosomiasis infection on the use of fibroscan for staging of fibrosis in chronic HCV patients | 2015 | Exclusion reason: Duplicate; |
| Serum hyaluronic acid as a non-invasive tool to diagnose schistosomal periportal fibrosis in Schistosoma mansoni endemic areas of Ethiopia | 2015 | Exclusion reason: Cannot source full text; authors did not respond; |
| Imported schistosomiasis in Italy: A single centre case series | 2015 | Exclusion reason: Duplicate; |
| Splenectomy improves hemostatic and liver functions in hepatosplenic schistosomiasis mansoni | 2015 | Exclusion reason: No comparator; |
| Schistosoma mansoni-Related Hepatosplenic Morbidity in Adult Population on Kome Island, Sengerema District, Tanzania | 2015 | Exclusion reason: Not enough information for outcome effect sizes; |
| Acute kidney injury in schistosomiasis: A retrospective cohort of 60 patients in Brazil | 2015 | Exclusion reason: No comparator; H |
| Malondialdehyde; lipid peroxidation plasma biomarker correlated with hepatic fibrosis in human Schistosoma mansoni infection | 2015 | Exclusion reason: No comparator; |
| AlteraÃ§Ãµes duodenais na hipertensÃ£o portal da esquistossomose mansÃ´nica | 2015 | Exclusion reason: Study not in English; |
| Role of fibroscan, TGF-beta1, and YKL-40 in the detection of hepatic fibrosis in chronic hepatitis C patients with and without schistosomiasis | 2015 | Exclusion reason: Wrong exposure diagnosis method; |
| Correlation between levels of liver fibrosis and liver fibrosis biochemical parameters of advanced schistosomiasis patients | 2014 | Exclusion reason: Study not in English; |
| Non-invasive assessment of liver fibrosis by fibroactin test in comparison to liver biopsy in chronic hepatitis c with and without schistosoma mansoni | 2014 | Exclusion reason: Wrong outcome measurement; |
| High Schistosoma mansoni disease burden in a rural district of western Zambia | 2014 | Exclusion reason: Not enough information for outcome effect sizes; |
| Correlation between platelet count and both liver fibrosis and spleen diameter in patients with Schistosomiasis mansoni | 2014 | Exclusion reason: No comparator; |
| Enhanced liver fibrosis (ELF) score for the evaluation of liver fibrosis in Schistosoma mansoni | 2014 | Exclusion reason: Cannot source full text; |
| Evaluation of portal hypertensive doppler parameters in patients with bilharzial periportal fibrosis | 2014 | Exclusion reason: Not enough information for outcome effect sizes; |
| Serum hyaluronic acid as a non-invasive tool to diagnose schistosomal periportal fibrosis in schistosoma mansoni endemic areas of Ethiopia | 2014 | Exclusion reason: Cannot source full text; authors did not respond; |
| Thrombocytopenia as a surrogate marker of hepatosplenic schistosomiasis in endemic areas for Schistosomiasis mansoni | 2014 | Exclusion reason: No comparator; |
| [Influence factors of Schistosoma japonicum infection among fishermen in eastern Dongting Lake Region] | 2013 | Exclusion reason: Study not in English; |
| A very high infection intensity of Schistosoma mansoni in a Ugandan Lake Victoria fishing community is required for association with highly prevalent organ related morbidity | 2013 | Exclusion reason: Not enough information for outcome effect sizes; |
| Treatment and education reduce the severity of schistosomiasis periportal fibrosis | 2013 | Exclusion reason: No comparator; |
| Micro-Geographical Heterogeneity in Schistosoma mansoni and S. haematobium Infection and Morbidity in a Co-Endemic Community in Northern Senegal | 2013 | Exclusion reason: Not enough information for outcome effect sizes; |
| Hemostatic dysfunction is increased in patients with hepatosplenic schistosomiasis mansoni and advanced periportal fibrosis | 2013 | Exclusion reason: Outcome not binary; |
| Relationship between splenomegaly and hematologic findings in patients with hepatosplenic schistosomiasis | 2013 | Exclusion reason: Not enough information for outcome effect sizes; |
| Fibroscan of chronic HCV patients coinfected with schistosomiasis | 2013 | Exclusion reason: Wrong outcome measurement; |
| Aspectos ultrassonogrÃ¡ficos associados Ã  morbidade de formas clÃ­nicas crÃ´nicas de esquistossomose mansÃ´nica, utilizando-se protocolo proposto pela OrganizaÃ§Ã£o Mundial da SaÃºde | 2013 | Exclusion reason: No comparator; |
| Role of CCR5DELTA32 mutation in protecting patients with Schistosoma mansoni infection against hepatitis C viral infection or progression | 2013 | Exclusion reason: No comparator; |
| Role of CCR532 mutation in protecting patients with Schistosoma mansoni infection against hepatitis C viral infection or progression | 2013 | Exclusion reason: No comparator; |
| Associating portal congestive gastropathy and hepatic fibrosis in hepatosplenic mansoni schistosomiasis | 2013 | Exclusion reason: No comparator; |
| Persistent organomegaly associated with schistosomiasis and correlates of abnormal liver pattern in young Kenyan children | 2013 | Exclusion reason: Duplicate; |
| Clinical and ultrasonographic correlates of hepatosplenic schistsomiasis among children and adults in a schistosoma mansoni hyperendemic rural area of zambia | 2013 | Exclusion reason: Not enough information for outcome effect sizes; |
| T lymphocyte profile and activation status in schistosomiasis patients with liver fibrosis | 2013 | Exclusion reason: Not enough information for outcome effect sizes; |
| Impaired lymphocyte profile in schistosomiasis patients with periportal fibrosis | 2013 | Exclusion reason: Not enough information for outcome effect sizes; |
| Immunomodulatory effect of R848 on cytokine production associated with Schistosoma mansoni infection | 2013 | Exclusion reason: No comparator; |
| Evaluation of pulmonary hypertension in Bilharzial patients | 2013 | Exclusion reason: Wrong exposure diagnosis method; |
| Hemorragia digestiva alta varicosa em hospital de emergÃªncia e, Recife - PE | 2013 | Exclusion reason: Study not in English; |
| Evaluation of portal hypertensive Doppler parameters in patients with periportal bilharzial hepatic fibrosis | 2013 | Exclusion reason: Duplicate; |
| Coinfection with hepatitis C virus and schistosomiasis: Fibrosis and treatment response | 2013 | Exclusion reason: Wrong outcome measurement; |
| Ultrasound study of liver disease caused by Schistosoma mansoni in rural Zambian schoolchildren | 2012 | Exclusion reason: Not enough information for outcome effect sizes; |
| Ultrasound and magnetic resonance imaging findings in Schistosomiasis mansoni: Expanded gallbladder fossa and fatty hilum signs | 2012 | Exclusion reason: Wrong exposure diagnosis method; |
| Comparison between transient elastography (Fibroscan) and liver biopsy for diagnosis of hepatic fibrosis in chronic hepatitis C genotype 4 | 2012 | Exclusion reason: Duplicate; |
| Serum and tissue osteopontin in human and murine schistosomiasis: Relationship with liver fibrosis | 2012 | Exclusion reason: Duplicate; |
| Association of MICA gene polymorphisms with liver fibrosis in schistosomiasis patients in the Dongting Lake region | 2012 | Exclusion reason: Outcome not binary; ; |
| Hepatosplenic morbidity due to Schistosoma mansoni in schoolchildren on Ukerewe Island, Tanzania | 2012 | Exclusion reason: Not enough information for outcome effect sizes; |
| Liver schistosomiasis and chronic hepatitis B: A deleterious association | 2012 | Exclusion reason: Duplicate; |
| Evaluation on intervention effect of a 17-year health promotion of schistosomiasis among residents in lake-type endemic area | 2011 | Exclusion reason: Cannot source full text; Cannot find this journal; |
| Aspectos epidemiolÃ³gicos da esquistossomose hepatoesplÃªnica no estado de Pernambuco, Brasil | 2011 | Exclusion reason: Study not in English; |
| Schistosomiasis mansoni: ultrasound-evaluated hepatic fibrosis and serum concentrations of hyaluronic acid | 2011 | Exclusion reason: Not enough information for outcome effect sizes; |
| Liver fibrosis and cirrhosis in hepatitis b and schistosomiasis co-infection category: Clinical lesson | 2011 | Exclusion reason: Not enough information for outcome effect sizes; |
| Nitric oxide production levels in patients with different stages of hepatosplenic schistosomiasis | 2011 | Exclusion reason: Cannot source full text; authors did not respond; |
| Access to antioxidant rich diet and development of schistosomal periportal fibrosis (PPF): Report from an Ethiopian cohort revisited after 10 years | 2011 | Exclusion reason: Duplicate; |
| Epidemiological evaluation of schistosomiasis in migrant fishermen in Dongting Lake region | 2010 | Exclusion reason: Study not in English; |
| Imaging techniques and histology in the evaluation of liver fibrosis in hepatosplenic schistosomiasis mansoni in Brazil: a comparative study | 2010 | Exclusion reason: No comparator; |
| Longitudinal observation on effect of health education for adult male residents for 18 years in heavy epidemic areas of schistosomiasis around Poyang Lake region | 2010 | Exclusion reason: Study not in English; |
| Schistosoma mansoni: magnetic resonance analysis of liver fibrosis according to WHO patterns for ultrasound assessment of schistosomiasis-related morbidity | 2010 | Exclusion reason: No comparator; |
| Factors controlling the effect of praziquantel on liver fibrosis in Schistosoma mansoni-infected patients | 2010 | Exclusion reason: No comparator; |
| Comparison between clinical and ultrasonographic findings in cases of periportal fibrosis in an endemic area for schistosomiasis mansoni in Brazil | 2010 | Exclusion reason: Not enough information for outcome effect sizes; |
| Serum hyaluronan and collagen IV as non-invasive markers of liver fibrosis in patients from an endemic area for schistosomiasis mansoni: a field-based study in Brazil | 2010 | Exclusion reason: Not enough information for outcome effect sizes; |
| Risk factors for bleeding in patients with asymptomatic oesophageal varices secondary to schistosomal portal hypertension: a longitudinal hospital based study | 2009 | Exclusion reason: Cannot source full text; Cannot find this journal; |
| Evaluation of fibrosis seromarkers versus liver biopsy in Egyptian patients with hepatitis C and/or NASH and/or schistosomiasis | 2009 | Exclusion reason: Cannot source full text; |
| TIMP-1 in response to egg antigens predicts hepatic fibrosis in human Schistosoma Japonicum infection | 2009 | Exclusion reason: Duplicate; |
| NÃ­veis sÃ©ricos de globulinas e a intensidade da fibrose hepÃ¡tica em pacientes com esquistossomose mansÃ´nica | 2009 | Exclusion reason: Study not in English; |
| Serum globulin levels and intensity of hepatic fibrosis in patients with mansonic schistosomiasis | 2009 | Exclusion reason: Study not in English; |
| Reversibility of schistosomal periportal thickening/fibrosis after praziquantel therapy: A twenty-six month follow-up study in Ethiopia | 2009 | Exclusion reason: Duplicate; |
| Effect of chemotherapy with praziquantel on the production of cytokines and morbidity associated with schistosomiasis mansoni | 2008 | Exclusion reason: No comparator; |
| Abdominal ultrasound in the evaluation of fibrosis and portal hypertension in an area of schistosomiasis low endemicity | 2008 | Exclusion reason: No comparator; |
| Detection of early liver fibrosis in patients with intestinal schistosomiasis: sonographic and histologic findings in Schistosoma mansoni infection | 2008 | Exclusion reason: Not enough information for outcome effect sizes; |
| Human schistosomiasis mansoni: immune responses during acute and chronic phases of the infection | 2008 | Exclusion reason: No comparator; |
| Reversibility of schistosomal periportal thickening/fibrosis after praziquantel therapy: a twenty-six month follow-up study in Ethiopia | 2008 | Exclusion reason: Outcome not binary; |
| AvaliaÃ§Ã£o da concordÃ¢ncia entre ressonÃ¢ncia magnÃ©tica de ultra-sonografia na classificaÃ§Ã£o de fibrose periportal em esquitossomÃ³ticos, segundo a classificaÃ§Ã£o de Niamey | 2007 | Exclusion reason: Study not in English; |
| Reprodutibilidade da classificaÃ§Ã£o ultra-sonogrÃ¡fica de Niamey na avaliaÃ§Ã£o da fibrose periportal na esquistossomose mansÃ´nica | 2007 | Exclusion reason: Study not in English; |
| Clinical and ultrasound findings before and after praziquantel treatment among Venezuelan schistosomiasis patients | 2007 | Exclusion reason: No comparator; |
| Reduced serum concentrations of retinol and a-tocopherol and high concentrations of hydroperoxides are associated with community levels of S. mansoni infection and schistosomal periportal fibrosis in Ethiopian school children | 2007 | Exclusion reason: Not enough information for outcome effect sizes; |
| Rule of schistosomiasis infection in chronic hepatitis C | 2007 | Exclusion reason: Wrong outcome measurement; |
| Liver morbidity due to Schistosoma mekongi in Cambodia after seven rounds of mass drug administration | 2007 | Exclusion reason: Wrong exposure; |
| Th2 cytokines are associated with persistent hepatic fibrosis in human Schistosoma japonicum infection | 2007 | Exclusion reason: Outcome not binary; |
| Reduced serum concentrations of retinol and alpha-tocopherol and high concentrations of hydroperoxides are associated with community levels of S. mansoni infection and schistosomal periportal fibrosis in Ethiopian school children | 2007 | Exclusion reason: Duplicate; |
| Clinical characteristics of schistosomiasis japonica liver fibrosis and its prophylaxis and treatment countermeasures | 2006 | Exclusion reason: Study not in English; |
| Schistosoma mansoni associated mortality in Gezira: Determined by clinical and ultrasound examination | 2006 | Exclusion reason: Cannot source full text; |
| Comparative analysis of ultrasonic evidence and serological findings of schistosomiasis liver fibrosis | 2006 | Exclusion reason: Study not in English; |
| Validity of inquiry in screening chronic schistosomiasis japonica | 2006 | Exclusion reason: Study not in English; |
| Treatment of hepatitis C virus genotype 4 with peginterferon alfa-2a: impact of bilharziasis and fibrosis stage | 2006 | Exclusion reason: Not enough information for outcome effect sizes; authors contacted; |
| Ultrasound and clinical investigation of hepatosplenic schistosomiasis: evaluation of splenomegaly and liver fibrosis four years after mass chemotherapy with oxamniquine | 2006 | Exclusion reason: Not enough information for outcome effect sizes; authors contacted; |
| Effects of repeated praziquantel treatment on schistosomiasis mekongi morbidity as detected by ultrasonography | 2006 | Exclusion reason: No comparator; |
| Large scale evaluation of WHO's ultrasonographic staging system of schistosomal periportal fibrosis in Ethiopia | 2006 | Exclusion reason: Not enough information for outcome effect sizes; |
| Clinical trial of spironolactone in treatment of hepatic fibrosis of schistosomiasis | 2005 | Exclusion reason: Study not in English; |
| Clinical study on the treatment of schistosomiasis hepatic fibrosis with vitamin E | 2005 | Exclusion reason: Study not in English; |
| Determination of serum levels of YKL-40 and hyaluronic acid in patients with hepatic fibrosis due to schistosomiasis japonica and appraisal of their clinical value | 2005 | Exclusion reason: Not enough information for outcome effect sizes; |
| Circulating adhesion molecules in patients with different clinical forms of S. mansoni infection | 2005 | Exclusion reason: No comparator; |
| Regression of liver fibrosis in Schistosoma mansoni infected Sudanese subjects after praziquantel treatment | 2005 | Exclusion reason: Editorial; |
| Analysis of type-B ultrasound of hepatic fibrosis for historical schistosomiasis patients | 2005 | Exclusion reason: Study not in English; |
| Role of non invasive biomarkers in the assessment of liver condition in chronic hepatitis C Egyptian patients and if they correlate with the severity of liver affection | 2005 | Exclusion reason: Cannot source full text; |
| Comparative clinical and ultrasound study of egg-negative and egg-positive individuals from Schistosoma mansoni low morbidity endemic areas, and hospitalized patients with hepatosplenic disease | 2005 | Exclusion reason: Not enough information for outcome effect sizes; |
| Urinary transforming growth factor beta 1 [TGF beta 1] and soluble intercellular adhesion molecule-1 [sICAM-1] as non invasive immunological indicators of disease progression and morbidity in human schistosomiasis mansoni and haematobium | 2005 | Exclusion reason: Cannot source full text; |
| Prevalence of schistomasiasis lesions detected by ultrasonography in children in Molodo, Mali | 2005 | Exclusion reason: Not enough information for outcome effect sizes; |
| Circulating markers of oxidative stress and liver fibrosis in Sudanese subjects at risk of schistosomiasis and hepatitis | 2005 | Exclusion reason: Wrong exposure; |
| Analysis on morbidity and chemotherapy effects of Schistosoma japonicum infection in fishermen on Dongting Lake | 2004 | Exclusion reason: Study not in English; |
| Detailed clinical and ultrasound examination of children and adolescents in a Schistosoma mansoni endemic area in Kenya: hepatosplenic disease in the absence of portal fibrosis | 2004 | Exclusion reason: No comparator; |
| Immunohistochemical detection of fas expression and Bcl2 in liver tissues of patients with chronic liver disease | 2004 | Exclusion reason: Wrong exposure diagnosis method; |
| Cytokine profile associated with human chronic schistosomiasis mansoni | 2004 | Exclusion reason: No comparator; |
| Effects of "Biejiaruangan" tablets in treatment of schistosomiasis cirrhosis | 2004 | Exclusion reason: Study not in English; |
| Hepatosplenic morbidity in two neighbouring communities in Uganda with high levels of Schistosoma mansori infection but very different durations of residence | 2004 | Exclusion reason: Duplicate; |
| Hepatosplenic morbidity in two neighbouring communities in Uganda with high levels of Schistosoma mansoni infection but very different durations of residence | 2004 | Exclusion reason: Not enough information for outcome effect sizes; |
| Periportal Fibrosis in Human Schistosoma mansoni Infection Is Associated with Low IL-10, Low IFN-gamma, High TNF-alpha, or Low RANTES, Depending on Age and Gender | 2004 | Exclusion reason: Not enough information for outcome effect sizes; |
| Sonographic evaluation of periportal fibrosis in children living in an endemic region for Schistosoma mansoni | 2004 | Exclusion reason: Study not in English; |
| [Sonographic evaluation of periportal fibrosis in children living in a Schistosoma mansoni endemic region] | 2004 | Exclusion reason: Duplicate; |
| Effects of HBV infection on hepatic fibrosis and level of Th1/Th2 cytokines in the patients with Schistosomiasis japonica | 2003 | Exclusion reason: Cannot source full text; |
| Identification of groups at high risk of severe schistosomal morbidity: a suggested plan for control | 2003 | Exclusion reason: Cannot source full text; |
| Detection of serum transforming growth factor beta 1 in diagnosis of hepatic fibrosis in schistosomiasis and evaluation of treatment efficacy | 2003 | Exclusion reason: Study not in English; |
| Gammaglutamyl transpeptidase activity in patients with schistosomiasis | 2003 | Exclusion reason: Not enough information for outcome effect sizes; |
| Long-term effect of single dose mass praziquantel chemotherapy on morbidity and mortality of S. mansoni in Gezira, Sudan | 2003 | Exclusion reason: Cannot source full text; |
| Duplex Doppler ultrasound of hepatic Schistosomiasis japonica: A study of 47 patients | 2003 | Exclusion reason: Wrong exposure; |
| Detection on the portal system haemodynamics of patients with chronic and advanced schistosomiasis and its clinical meaning | 2002 | Exclusion reason: Cannot source full text; |
| Five-year impact of repeated praziquantel treatment on subclinical morbidity due to Schistosoma japonicum in China | 2002 | Exclusion reason: Wrong exposure; |
| Cytokine regulation of periportal fibrosis in humans infected with Schistosoma mansoni: IFN-gamma is associated with protection against fibrosis and TNF-alpha with aggravation of disease | 2002 | Exclusion reason: Not enough information for outcome effect sizes; |
| Study on histopathology, ultrasonography and some special serum enzymes and collagens for 38 advanced patients of schistosomiasis japonica | 2002 | Exclusion reason: No comparator; |
| Hepatosplenic schistosomiasis in field-based studies: a combined clinical and sonographic definition | 2001 | Exclusion reason: Wrong exposure; |
| Hepatosplenic schistosomiasis in field-based studies: a combined clinical and sonographic definition | 2001 | Exclusion reason: Duplicate; |
| Biliary disorders in patients with schistosomal hepatic fibrosis | 2001 | Exclusion reason: Cannot source full text; |
| Quantitative evaluation and its significance of hepatic fibrosis in patients with advanced schistosomiasis | 2000 | Exclusion reason: Cannot source full text; |
| Observation on changes of liver fibrosis due to Schistosoma japonicum after treatment with praziquantel | 2000 | Exclusion reason: Study not in English; |
| Hyaluronate levels and markers of oxidative stress in the serum of Sudanese subjects at risk of infection with Schistosoma mansoni | 2000 | Exclusion reason: Wrong exposure; |
| Results of sclerotherapy for bleeding esophageal varices in patients with schistosomal liver disease. A retrospective study | 2000 | Exclusion reason: Cannot source full text; |
| Effects of single-dose praziquantel on morbidity and mortality resulting from intestinal schistosomiasis | 2000 | Exclusion reason: Not enough information for outcome effect sizes; |
| Two-year impact of praziquantel treatment for Schistosoma japonicum infection in China: re-infection, subclinical disease and fibrosis marker measurements | 2000 | Exclusion reason: Not enough information for outcome effect sizes; |
| Hepatic fibrosis due to fascioliasis and/or schistosomiasis in Abis 1 village, Egypt | 2000 | Exclusion reason: Not enough information for outcome effect sizes; |
| Clinical, virological and histopathological features: long-term follow-up in patients with chronic hepatitis C co-infected with S. mansoni | 2000 | Exclusion reason: Wrong outcome measurement; |
| Morbidity associated with Schistosoma mansoni infection determined by ultrasound in an endemic area of Brazil Caatinga do Moura | 2000 | Exclusion reason: No comparator; |
| The Epidemiology of Schistosomiasis in Egypt: qena Governorate | 2000 | Exclusion reason: Schistosoma haematobium; |
| The epidemiology of schistosomiasis in Egypt: Assiut governorate | 2000 | Exclusion reason: Schistosoma haematobium; |
| The epidemiology of schistosomiasis in Egypt: Minya Governorate | 2000 | Exclusion reason: Schistosoma haematobium; |
| The epidemiology of schistosomiasis in Egypt: summary findings in nine governorates | 2000 | Exclusion reason: Report; |
| The epidemiology of schistosomiasis in Egypt: fayoum Governorate | 2000 | Exclusion reason: Not enough information for outcome effect sizes; |
| Use of a Tablet-Based System to Perform Abdominal Ultrasounds in a Field Investigation of Schistosomiasis-Related Morbidity in Western Kenya |  | Exclusion reason: Editorial; |
| Schistosomiasis in immigrants, refugees and travellers in an Italian referral centre for tropical diseases |  | Exclusion reason: Not enough information for outcome effect sizes; |
| Egy-Score Can Predict Portal Hypertension and Oesophageal Varices in Chronic Hepatitis C with Good Sensitivity, Specificity and Diagnostic Accuracy |  | Exclusion reason: Wrong outcomes; |
| Chronic Hepatosplenomegaly in African School Children: A Common but Neglected Morbidity Associated with Schistosomiasis and Malaria |  | Exclusion reason: Editorial; |
| Prevalence of schistosome antibodies with hepatosplenic signs and symptoms among patients from Kaoma, Western Province, Zambia |  | Exclusion reason: Wrong outcomes; |
| Chronic hepatitis B and liver schistosomiasis: a deleterious association |  | Exclusion reason: Not enough information for outcome effect sizes; |
| Risk of hepatitis "E" virus infection among some schistosomiasis patients in Egypt | 1995 | Exclusion reason: Wrong outcomes; |
| Sonographical morphometrical findings of the liver and spleen in Sudanese patients with Schistosoma mansoni induced periportal fibrosis | 1994 | Exclusion reason: Outcome not binary; |
| Characteristic sonographic pattern of schistosomal hepatic fibrosis | 1989 | Exclusion reason: Wrong exposure diagnosis method; |
| Prevalence of schistosoma mansoni and intestinal parasites with evaluation of hepatic schistosomiasis in a rural area after governmental efforts (Gharbia governorate) | 1997 | Exclusion reason: Cannot source full text; Inter-Library Request (number 5013896); |
| Pulmonary hypertension in schistosomiasis mansoni | 1996 | Exclusion reason: Not enough information for outcome effect sizes; |
| Effect of the Benzthiadiazines on the Portal Pressure in Schistosomal Hepatic Fibrosis | 1963 | Exclusion reason: Outcome not binary; |
| Effect of splenectomy on the portal pressure in bilharzial hepatic fibrosis | 1962 | Exclusion reason: Report; |
| Reversibility of Schistosoma mansoni-associated morbidity after yearly mass praziquantel therapy: ultrasonographic assessment | 1998 | Exclusion reason: "After" in before-after study; |
| Changes of ultrasonography and two serum biochemical indices for hepatic fibrosis in schistosomiasis japonica patients one year after praziquantel treatment | 1997 | Exclusion reason: Not enough information for outcome effect sizes; |
| Ultrasonographical investigation of periportal fibrosis in children with Schistosoma mansoni infection: reversibility of morbidity twenty-three months after treatment with praziquantel | 1992 | Exclusion reason: Duplicate; |
| [A new diuretic: Furosemide. Its effect on bilahariasis-cirrhosis with ascites] | 1965 | Exclusion reason: Study not in English; |
| Assessment of liver function by the MEGX test in patients with schistosomiasis and cirrhosis | 1999 | Exclusion reason: Wrong exposure diagnosis method; |
| Trial of hydrochlorothiazide in ascites of bilharzial hepatolienal fibrosis | 1962 | Exclusion reason: Outcome not binary; |
| Prevalence and morbidity of schistosomiasis among rural fishermen at two Egyptian villages (Gharbia Governorate) | 1995 | Exclusion reason: Not enough information for outcome effect sizes; |
| Serum growth hormone and somatomedin-C in patients with bilharzial hepatic fibrosis and post hepatitic cirrhosis | 1991 | Exclusion reason: Not enough information for outcome effect sizes; |
| Correlation between the level of serum growth hormone; somatomedin-C; anthropometric measurements and the liver functions in chronic liver diseases | 1991 | Exclusion reason: Not enough information for outcome effect sizes; |
| The pharmacokinetics of antipyrine in patients with graded severity of schistosomiasis | 1985 | Exclusion reason: Wrong outcome measurement; |
| Evidence for a long-term effect of a single dose of praziquantel on Schistosoma mansoni-induced hepatosplenic lesions in northern Uganda | 1999 | Exclusion reason: Not enough information for outcome effect sizes; |
| Ultrasonography of periportal fibrosis in schistosomiasis mansoni in Brazil | 1997 | Exclusion reason: Not enough information for outcome effect sizes; |
| Hepatitis B vaccination in children infected with Schistosoma mansoni: correlation with ultrasonographic data | 1990 | Exclusion reason: Wrong outcomes; |
| Immunogenetic susceptibility for post-schistosomal hepatic fibrosis | 1991 | Exclusion reason: No comparator; |
| Effect of doxycycline administration on nitrogen excretion in bilharzial hepatic fibrosis and chronic renal failure cases | 1976 | Exclusion reason: Wrong outcome measurement; |
| Diagnosis of pathologically confirmed Symmers' periportal fibrosis by ultrasonography: a prospective blinded study | 1988 | Exclusion reason: Outcome not binary; |
| Association of the therapeutic activity of praziquantel with the reversal of Symmers' fibrosis induced by Schistosoma mansoni | 1991 | Exclusion reason: "After" in before-after study; |
| Effect of antischistosomal chemotherapy on prevalence of Symmers' periportal fibrosis in Sudanese villages | 1988 | Exclusion reason: Duplicate; |
| Efficacy and tolerance of praziquantel in patients with Schistosoma mansoni infection and Symmers' fibrosis: a field study in the Sudan | 1988 | Exclusion reason: Wrong outcome measurement; |
| The effectiveness of annual versus biennial mass chemotherapy in reducing morbidity due to schistosomiasis: a prospective study in Gezira-Managil, Sudan | 1996 | Exclusion reason: Not enough information for outcome effect sizes; |
| Use of ultrasound in a study of schistosomal periportal fibrosis in rural Zimbabwe | 1993 | Exclusion reason: Wrong exposure; |
| Schistosoma mansoni-related morbidity on Ukerewe Island, Tanzania: clinical, ultrasonographical and biochemical parameters | 1997 | Exclusion reason: Not enough information for outcome effect sizes; |
| Hepatosplenic morbidity in schistosomiasis japonica: evaluation with Doppler sonography | 1999 | Exclusion reason: Not enough information for outcome effect sizes; |
| Mortality due to schistosomiasis mansoni: a field study in Sudan | 1999 | Exclusion reason: "After" in before-after study; |
| [Value of echography in the study of periportal fibrosis caused by schistosomiasis in an endemic African zone] | 1990 | Exclusion reason: Study not in English; |
| Impact mass chemotherapy with praziquantel on schistosomiasis control in Fanhu village, People's Republic of China | 1997 | Exclusion reason: "After" in before-after study; |
| [The morbidity investigation of residents in a highly endemic village of schistosomiasis in Poyang Lake region] | 1998 | Exclusion reason: Study not in English; |
| Chronic liver disease in black children in Durban, South Africa | 1984 | Exclusion reason: Wrong exposure diagnosis method; |
| [Functional and immunologic evaluation of prothrombin in the hepatosplenic form of schistosomiasis and hepatic cirrhosis] | 1983 | Exclusion reason: Study not in English; |
| Ultrasonographical investigation of periportal fibrosis in children with Schistosoma mansoni infection: reversibility of morbidity seven months after treatment with praziquantel | 1991 | Exclusion reason: No comparator; |
| [Periportal fibrosis in Schistosoma mansoni schistosomiasis. An ultrasonographic study in the area of Nkolbisson (Cameroon)] | 1993 | Exclusion reason: Study not in English; |
| Relationships between several markers of extracellular matrix turn-over and ultrasonography in human Schistosomiasis mansoni | 1999 | Exclusion reason: Wrong outcome measurement; |
| Hepatosplenic schistosomiasis: comparison of sonographic findings in Brazilian and Sudanese patients--correlation of sonographic findings with clinical symptoms | 1992 | Exclusion reason: Not enough information for outcome effect sizes; |
| Epidemiological identification of Chinese individuals putatively susceptible or insusceptible to Schistosoma japonicum: a prelude to immunogenetic study of human resistance to Asian schistosomiasis | 1998 | Exclusion reason: Not enough information for outcome effect sizes; |
| Clinical, parasitological and immunological features of canal cleaners hyper-exposed to Schistosoma mansoni in the Sudan | 1996 | Exclusion reason: Outcome not binary; |
| Radioactive chromium studies in bilharzial hepatic fibrosis and splenomegaly before and after splenectomy | 1972 | Exclusion reason: Outcome not binary; |
| Schistosomiasis japonica on Jishan Island, Jiangxi Province, People's Republic of China: persistence of hepatic fibrosis after reduction of the prevalence of infection with age | 1993 | Exclusion reason: Not enough information for outcome effect sizes; |
| Impact of annual screening and chemotherapy with praziquantel on schistosomiasis japonica on Jishan Island, People's Republic of China | 1994 | Exclusion reason: "After" in before-after study; |
| [Determination of the thickness of the wall of portal vein trunk in patients of schistosomiasis japonica with hepatic fibrosis and its clinical significance] | 1994 | Exclusion reason: Study not in English; |
| Could we diagnose active schistosomiasis in children by abdominal sonography? | 1996 | Exclusion reason: Report; |
| Why compare, with the help of echography, the morbidity of hepatosplenic schistosomiasis in six African countries? Presentation of a WHO project that includes Madagascar | 1994 | Exclusion reason: Study not in English; |
| Effect of antischistosomal chemotherapy on prevalence of Symmers' periportal fibrosis in Sudanese villages | 1988 | Exclusion reason: Not enough information for outcome effect sizes; |
| Evaluation of liver fibrosis in Schistosoma japonicum infections using B-ultrasonic scan and serum hyaluronidase level | 1996 | Exclusion reason: Study not in English; |
| Effectiveness of selective chemotherapy in islet-circle marsh subtype endemic area of schistosomiasis in Poyang Lake | 1997 | Exclusion reason: Study not in English; |
| Queixadinha Project: morbidity and control of schistosomiasis in an endemic area in the northeast of the state of Minas Gerais, Brazil | 1996 | Exclusion reason: Study not in English; |
| A longitudinal study on morbidity control of schistosomiasis by mass chemotherapy | 1996 | Exclusion reason: Study not in English; |
| Sonographic organometry in Brazilian and Sudanese patients with hepatosplenic schistosomiasis mansoni and its relation to the risk of bleeding from oesophageal varices | 1992 | Exclusion reason: Wrong outcome measurement; |
| Retrospective ultrasonographic study of gall bladder changes in patients with common liver diseases in Egypt | 1993 | Exclusion reason: Wrong exposure; |
| Serum zinc and vitamin A levels in Egyptian patients with schistosomal and non-schistosomal chronic liver disease | 1990 | Exclusion reason: Wrong exposure; |
| Longitudinal study on schistosomiasis control in lake regions by mass chemotherapy combined with health education | 1997 | Exclusion reason: Study not in English; |
| A comparative study between serological and ultrasound diagnosis in schistosomiasis japonica liver fibrosis | 1999 | Exclusion reason: Study not in English; |
| Phenotypic characterization of CD4<sup>+</sup> T lymphocytes in periportal fibrosis secondary to schistosomiasis | 2021 | Exclusion reason: No comparator; |
| The epidemiology of schistosomiasis in Egypt: Patterns of Schistosoma mansoni infection and morbidity in Kafr El-Sheikh | 2000 | Exclusion reason: Duplicate; |
| PREVALENCE AND CLINICAL SIGNIFICANCE OF SCHISTOSOMIASIS-CHRONIC HEPATITIS B VIRUS CO-INFECTION IN ZAMBIA | 2017 | Exclusion reason: Wrong exposure; |
| Queixadinha Project: Morbidity and control of schistosomiasis in an endemic area in the northeast of State of Minas Gerais, Brazil | 1996 | Exclusion reason: Study not in English; |
| Morbidity due to schistosiomiasis japonica in the People's Republic of China | 1992 | Exclusion reason: "After" in before-after study; |
| Study of schistosomiasis among patients attending Hossinea Hospital, Sharkia Governorate | 1985 | Exclusion reason: Wrong outcomes; |
| <The> role of histamine, serotonin and catecholamines in portal hypertension related to Egyptian hepatic schistosomiasis | 1984 | Exclusion reason: Outcome not binary; |
| Gall bladder wall thickness: an indirect sonographic sign of portal hypertension in schistosomal hepatic fibrosis | 1990 | Exclusion reason: Report; |
| Serum fatty acid changes in typhoid fever and bilharzial hepatic fibrosis | 1988 | Exclusion reason: Wrong outcomes; |
| Hepatosplenic schistosomiasis associated with schistosoma mansoni infection in Yemenis living in Riyadh Saudi Arabia | 1994 | Exclusion reason: Cannot source full text; |
| <The> value of ultrasonography in investigation of morbidity of Schistosoma mansoni in Shebin El Kom, Menoufia governorate, Egypt | 1991 | Exclusion reason: Cannot source full text; |
| <The> role of ultrasonography to discriminate between schistosomiasis and virus-related chronic liver disease | 1999 | Exclusion reason: Cannot source full text; |
| Ultrasonographic and portal hemodynamic changes in hepatic schistosomiasis patients after treatment with praziquantel | 1994 | Exclusion reason: Cannot source full text; |
| Ultrasonographic and portal hemodynamic changes in hepatic schistosomiasis patients after treatment with praziquantel | 1995 | Exclusion reason: Cannot source full text; |
| Correlation of lesser omental thickness with portal venous pressure in portal hypertension secondary to schistosomal hepatic fibrosis | 1990 | Exclusion reason: Cannot source full text; |
| O Projeto Queixadinha: a morbidade e o controle da esquistossomose em Ã¡rea endÃªmica no nordeste de Minas Gerais, Brasil | 1996 | Exclusion reason: Study not in English; |
| Serum insulin and blood glucose levels during intravenous glucose tolerance test [IVGTT] in bilharzial and post-hepatitis cirrhosis | 1988 | Exclusion reason: Cannot source full text; |
| Correlation between the degree of periportal thickening, portal blood flow and splenic volume | 1997 | Exclusion reason: No comparator; |
| Study of fibronectin in patients with bilharzial hepatic fibrosis | 1989 | Exclusion reason: Cannot source full text; |
| Endoscopia de urgencia na hemorragia digestiva alta em pacientes esquistossomoticos ou cirroticos com hipertensao portal | 1982 | Exclusion reason: Study not in English; |
| Serum bile acids in schistosomotic hepatic fibrosis | 1992 | Exclusion reason: Report; |
| Immunoglobulin: a and pathogenesis of schistosomal glomerulopathy | 1995 | Exclusion reason: Wrong outcome measurement; |
| Evaluation of renal arterial blood flow in patients with various stages of schistosomal hepatic fibrosis by duplex ultrasonography | 1995 | Exclusion reason: Cannot source full text; |
| ContribuiÃ§Ã¤o ao estudo clÃ­nico e eletrencefalogrÃ¡fico da encefalopatia hepÃ¡tica nas cirroses e na esquistossomose mansÃ´nica | 1983 | Exclusion reason: Study not in English; |
| Grading of schistosomiasis patients' hepatic lesions by ultrasonography and analysis of portal vein and splenic vein haemodynamics | 1989 | Exclusion reason: Study not in English; |
| Comparative analysis of ultrasonic evidences and serological findings of schistosomiasis liver fibrosis | 1989 | Exclusion reason: Study not in English; |
| Clinical characteristics of schistosomiasis japonica liver fibrosis and its prophylaxis and treatment countermeasures | 1992 | Exclusion reason: Study not in English; |
| Doppler sonography of the portal circulation in cases with portal hypertension | 1997 | Exclusion reason: Cannot source full text; |

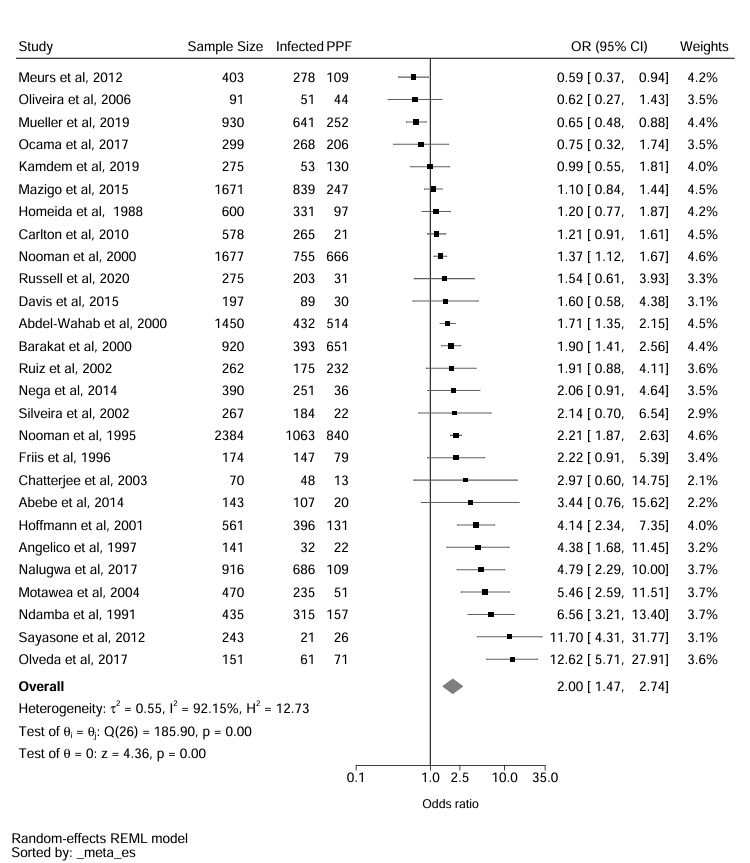
Figure S1. Pooled Forrest plot with outliers removed

*The three studies were Rouquet et al, 1993., Doehring-Schwerdtfeger et al, 1990., and Hassan et al, 1999*

|  | **With outliers** | | | | **With outliers removed** | | | |
| --- | --- | --- | --- | --- | --- | --- | --- | --- |
|  | **Number of studies** | **OR (95% CI)** | **I^2^** | **Test of group differences** | **Number of studies** | **OR (95% CI)** | **I^2^** | **Test of group differences** |
| **Continent** |  |  |  | Q_b_(2) = 3·07, p = 0·21 |  |  |  | Q_b_(2) = 2.47, p = 0·29 |
| Africa | 24 | 2·23 (1·51-3·30) | 94·28% |  | 21 | 1.84 (1.35-2.50) | 90.43% |  |
| Asia | 3 | 5·35 (1·13-25·40) | 93·80% |  | 3 | 5·35 (1·13-25·40) | 93·80% |  |
| South America | 3 | 1·32 (0·60-2·92) | 57·47% |  | 3 | 1·32 (0·60-2·92) | 57·47% |  |
| **Species** |  |  |  | Q_b_(1) = 1·09, p = 0·30 |  |  |  | Q_b_(1) = 1·87, p = 0·17 |
| *S. mansoni* | 27 | 2·27 (1·56-3·32) | 94·07% |  | 24 | 1.77 (1.33-2.35) | 88.97 |  |
| *S. japonicum/S. mekongi* | 3 | 5·35 (1·13-25·40) | 93·80% |  | 3 | 5·35 (1·13-25·40) | 93·80% |  |
| **Study setting** |  |  |  | Q_b_(2) =0·51 , p = 0·78 |  |  |  | Q_b_(2) =0·08, p = 0·96 |
| Community | 20 | 2·27 (1·42-3·62) | 96·40% |  | 18 | 1.74 (1.24-2.46) | 93.05% |  |
| School | 4 | 3·39 (0·81-14·13) | 89·12% |  | 3 | 2.02 (0.72-5.64) | 82.25% |  |
| Health Clinic | 4 | 1·86 (0·81-4·31) | 60·29% |  | 4 | 1·86 (0·81-4·31) | 60·29% |  |
| **Study Design** |  |  |  | Q_b_(1) = 15·75, p < 0· 001* |  |  |  |  |
| Cross-sectional | 25 | 1·83 (1·34-2·49) | 91·60% |  | 25 | Subgroup analyses not performed as less than three studies remaining in this group | | |
| Case-control | 5 | 15·80 (5·70-43·82) | 76·84% |  | 2 |  |  |  |
| **Sub-Saharan Africa MDA** |  |  |  | Q_b_(1) = 9·90, p < 0· 001* |  |  |  | Q_b_(1) = 5.73 ,  p < 0·02 |
| Pre-MDA (pre-2003) | 14 | 3·93 (2·23-6·94) | 95·99% |  | 11 | 2.45 (1.74 – 3.45) | 88.49% |  |
| Post-MDA (2003 and after) | 10 | 1·25 (0·82-1·92) | 81·41% |  | 10 | 1·25 (0·82-1·92) | 81·41% |  |
| **Protocol** |  |  |  | Q_b_(2) = 2·05, p = 0·36 |  |  |  | Q_b_(2) = 0.38, p = 0·83 |
| Niamey | 11 | 1·63 (0·91-2·94) | 90·52% |  | 11 | 1·63 (0·91-2·94) | 90·52% |  |
| Cairo | 9 | 1·94 (1·49-2·53) | 81·29% |  | 9 | 1·94 (1·49-2·53) | 81·29% |  |
| Alternative | 7 | 4·19 (1·33-13·23) | 92·46% |  | 5 | 2.14 (0.93 – 4.94) | 86.81% |  |
| **Niamey Deviations** |  |  |  | Q_b_(1) = 13·90, p = 0·01*** |  |  |  |  |
| Niamey | 4 | 3·13 (1·67-5·86) | 22·37% |  | None of the outliers used the Niamey protocol. Therefore, this subgroup remained unchanged | | | |
| Deviations from Niamey | 5 | 0·82 (0·59-1·13) | 59·16% |  |  |  |  |  |
| **Risk of Bias** |  |  |  | Q_b_(2) = 3·05, p = 0·22 |  |  |  | Q_b_(2) = 0.74, p = 0·69 |
| Low | 7 | 2·71 (0·98-7·48) | 95·55% |  | 6 | 1.70 (0.83 -3.47) | 89.77 |  |
| Medium | 16 | 1·92 (1·34-2·76) | 92·41% |  | 16 | 1·92 (1·34-2·76) | 92·41% |  |
| High | 7 | 5·53 (1·71-17·87) | 87·32% |  | 5 | 2.91 (1.06-7.99) | 83.32% |  |

Table S10: Subgroup analyses with the three outlier studies removed

*The three studies were Rouquet et al, 1993., Doehring-Schwerdtfeger et al, 1990., and Hassan et al, 1999*

Text S3: Infection category analyses

Three out of four studies that reported adjusted odds ratios used a low infection group (1-99 *S. mansoni* eggs per gram of stool) as the reference group.^21,31,35^ A single study by, Davis*, p*resented the odds ratios for both a low infection group as a comparator and uninfected as a comparator, the low reference group was used to calculate the pooled effect to allow comparison^.21^ For infection intensity, four studies reported adjusted effect sizes using WHO intensity categories;^6^ of these studies were the only analyses that could be meta-analysed.^21,31,35,50^ Three studies adjusted effect sizes based on infection intensity but could not be combined due to unique intensity classifications.^29,39,43^

**Table S10: Full Risk of Bias analyses**

| **Study ID** | **D1** | **D2** | **D3** | **D4** | **D5** | **D6** | **D7** | **D8** | **D9** | **D10** | **D11** | **D12** | **D13** | **D14** | **Score** | **Risk of bias** |
| --- | --- | --- | --- | --- | --- | --- | --- | --- | --- | --- | --- | --- | --- | --- | --- | --- |
| Abdel-Wahab et al, 2000 | 1, yes. Specific aim and analyses match what is reported | 1, yes. Protocol for study described the study population of interest. | 1, yes. Stratified random sampling applied | 0, no. No information given about non-responders or differences between known characteristics | 1, yes. Sample was representative of specified population | 1, yes. Justification of sample size given in study design paper | 0, unclear. Does not explicitly say the KK* was done before USS** | 1, yes. Infection intensity defined and standardised microscopy measurement techniques. | N/A | 0, no. Does not explicitly say that two USS measurement were taken but they followed the Cairo protocol | 1, yes. Microscopy = parasitologist, USS = trained physicians | 1, yes. 8.3% did not provide stool samples | 1, yes. Risk factors defined and categorised. | 0, unclear. Was not informed of whether the effect measures were adjusted or unadjusted. | 9 | M |
| Abebe et al, 2014^38^ | 1, yes. Specific aim and analyses match what is reported | 1, yes. Residents of Alamata District. Time period given (Dec 2011- Mar 2012) | 1, yes. Simple random sampling of all 1051 households | 0, no. No information given about non-responders or differences between known characteristics | 1, yes. Random sample from the community. | 1, yes. Sample size calculation is given. | 1, yes. KK was performed before USS. | 1, yes. Infection intensity categorised and 10% of the slides were re-checked | N/A | 1, yes. Two measurements taken (image pattern and portal branch wall thickness) | 1, yes. Microscopy = technicians, USS= clinicians | 0, unclear. Does not report whether there is missing data. | 1, yes. Only risk factor was age in years: 1-10, 10.1-20.0, 20.1-30.0, 30.0+ | 0, no. Descriptive statistics only | 10 | L |
| Angelico et al, 1997 | 1. yes. Specific aim and analyses match what is reported | 1, yes. Noted location, inclusion criteria, demographic table, and study year | 1, yes. Stated opportunistic sampling strategy. Detailed information provided about the demographics. | 0, no. No details on the non-respondents or wider demographic | 0, no. Sample only included those who had suspected chronic liver disease. Exclusion criteria not stated. | 0, no. No power calculations or justification | 0, no. Sample was recruited based on liver disease status then tested for schisto | 1, yes. Multiple eligible methods of detecting infection | 0, unclear. No mention of who performed KK/USS and their knowledge of the participants infection status | 1, yes. Cited Cairo method, at least 6 measurements taken. No mention of trained status but conducted at a hospital | 0, unclear. No mention who performed KK/USS | 0, no. Missing participant data. No model presented | 1, yes. Only adults and the covariates of rural and gender was consistent throughout the study. | 0, no. No model for PPF | 6 | M |
| **Study ID** | **D1** | **D2** | **D3** | **D4** | **D5** | **D6** | **D7** | **D8** | **D9** | **D10** | **D11** | **D12** | **D13** | **D14** | **Score** | **Risk of bias** |
| Barakat et al, 2000^39^ | 1, yes. Specific aim and analyses match what is reported. | 1, yes. Protocol for study described the study population of interest. | 1, yes. Stratified random sampling applied | 0, no. No information given about non-responders or differences between known characteristics | 1, yes. Sample was representative of specified population | 1, yes. Justification of sample size given in study design paper | 0, unclear. Does not explicitly say the KK was done before USS | 1, yes. Infection intensity defined and standardised microscopy measurement techniques. | N/A | 0, no. Does not explicitly say that two USS measurement were taken but they followed the Cairo protocol | 1, yes. Microscopy = technicians, USS= clinicians | 0, unclear. Does not report who is missing from the study | 1, yes. Risk factors defined and categorised. | 0, unclear. Does not describe how the effect measures were obtained | 8 | M |
| Carlton et al, 2010^40^ | 1, yes. Specific aim and analyses match what is reported. | 1, yes. Participants were from selected from a high infection prevalence sample. | 1, yes. Stratified random sampling. | 0, no. No information given about non-responders or differences between known characteristics. | 1, yes. Clear inclusion criteria described and representative of the underlying sample. | 0, no. Sample size justification is not given. | 1, yes. Parasitic investigation done before USS. | 1, yes. Infection status is defined by the positivity of either the miracidial or KK test. Infection intensity in eggs per gram but divided into quartiles. | N/A | 0, no. Does not explicitly say that two USS measurement were taken | 0, unclear. Does not state who measured infection status | 0,no. 28% of study lost to follow up. | 1, yes. Covariates were age, sex, occupation, educational attainment, and previous infection status. | 1, yes. Unadjusted and adjusted ORs presented using generalised estimating logistic regression model | 8 | M |
| Chatterjee et al, 2003^41^ | 1, yes. Specific aim and analyses match what is reported | 1, yes. Location, time period, and participant characteristics in terms of their infection and morbidity status | 0, no. No details about sampling method | 0, no. No information given about non-responders or differences between known characteristics | 0, no. No clear eligibility criteria. | 0, no. No sample size justification provided. | 0, unclear. Does not explicitly say the KK was done before USS | 1, yes. Infection status/intensity measured in two samples. | N/A | 0, no. Does not state that two USS measurements were taken. | 0, unclear. Does not state who measured infection status or PPF | 1, yes. 15/85 of the participants were not examined for PPF. | 0, unclear | 0, no. Descriptive statistics only | 4 | H |
| **Study ID** | **D1** | **D2** | **D3** | **D4** | **D5** | **D6** | **D7** | **D8** | **D9** | **D10** | **D11** | **D12** | **D13** | **D14** | **Score** | **Risk of bias** |
| Davis et al, 2015^32^ | 1, yes. Specific aim and analyses match what is reported. | 1, yes. Participants selected from villages with a high prevalence of  *S. mansoni* | 1, yes. Convenience sampling. | 0, no. No information given about non-responders.  The differences between known characteristics are provided but without statistical assessment | 0, unclear. Clear eligibility criteria but study uses convenience sampling reduces representativeness. | 1, yes. Justification of the sample size is given. Small due to lack for resources. | 0, unclear. Does not explicitly say the KK was done before USS | 1, yes. Infection status defined as the presence of eggs in stool. Infection intensity categorised using WHO categories. Quality assurance via an independent microscopist | N/A | 1, yes. Multiple images and measurements taken with an expert review. | 1, yes. Microscopy = microscopist, USS= sonographers | 1, yes. Only 2% of full data for children. | 1, yes. All variable defined. | 1, yes. Both adjusted and unadjusted prevalence risk ratios presented. | 10 | L |
| Doehring-Schwerdtfeger et al, 1990 | 1. yes. Specific aim and analyses match what is reported | 0, no. No study year | 0, no. Did not provide details about the how the sample was selected | 0, no. No details on the non-respondents or wider demographic | 1, yes. Controls were matched from the same school based upon age. | 0, no. No power calculations or justification | 1, yes. Infection status determined before the sonography | 0, unclear. No mention of who performed the microscopy or the quality control | 0, unclear. Did not state who performed KK/USS and their knowledge of the participants infection status | 0, unclear. Used a non-validated protocol. No mention of the training of the sonographer | 0, unclear. No mention who performed KK/USS | 0, not applicable. No model to include data in and there is no follow up | 1, yes. Infection intensity remains consistent | 0, no. No model | 4 | H |
| Friis et al, 1996^42^ | 1, yes. Specific aim and analyses match what is reported. | 0, no. School-attendees but time-period not given. | 1, yes. Attending one of the primary schools and between Grade 3-6. | 1, yes. 55.6% of the children were assessed. | 1, yes. All children from the same sampling frame. | 0, no. Does not state why these school children were targeted or if the sample is large enough for statistical power | 0, unclear. Does not explicitly say the KK was done before USS. | 1, yes. Defines infection status and intensity and a median egg count is taken over three days. | N/A | 1, yes. Three measurements were taken by a trained sonographer | 0, unclear. Does not state who measured infection status | 1, yes. All 174 children present in analysis | 1, yes. Covariates defined. | 0, no. Descriptive statistics only. | 8 | M |
| **Study ID** | **D1** | **D2** | **D3** | **D4** | **D5** | **D6** | **D7** | **D8** | **D9** | **D10** | **D11** | **D12** | **D13** | **D14** | **Score** | **Risk of bias** |
| Hassan et al 1999 | 1, yes. Specific aim and analyses match what is reported. | 0, no. No study year | 1, yes. Age matched negative controls and non-schistosomiasis helminth infection controls | 0, no. No details on the non-respondents/wider demographic | 1, yes. Cases and controls were from the same population and matched for age. | 0, no. No power calculations or justification | 1, yes. Stool and urine samples submitted and screened prior to the study start. | 0, unclear. No quality control and a new method was being tested which found low agreement with the egg counts | 0, unclear. No mention of who performed KK/USS and their knowledge of the status | 0, no. No validated methodology, but trained sonographer and more than 2 measurements | 0, unclear. No mention of who conducted the microscopy but there was a trained sonographer | 0, No. Missing data only included the 98 *S. mansoni* infected participants, no information on controls | 0, No. No detailed covariate information was provided. | 0, No. No model | 4 | H |
| Hirata et al, 1988 | 1. yes. Specific aim and analyses match what is reported | 0, no. No study year | 1, yes. Samples were list of previous patients, and all invited to participate in the study | 0, no. No details of the original number of people on the health registry | 0, unclear. No details of the community and the numbers of non-respondents | 0, no. No power calculations or justification | 1, Yes. All participants had been infected with schistosomiasis years earlier but not actively infected | 0, no. The study did not measure current infection. Used serology to determine chronic infection in a previously endemic area. | 0, unclear. No mention of who performed KK/USS and their knowledge of the participants infection status | 0, no. Insufficient detail of the ultrasound protocol | 0, unclear. No mention of who conducted serology/US | 0, not applicable. No model to include data in and there is no follow up | 0, no. No covariate information was provided (no age/gender/location information) | 0, no. No model | 3 | H |
| Homedia et al, 1988 | 1, yes. Specific aim and analyses match what is reported. | 0, no. No study year | 1, Yes. Random sample of a census | 0, no. Does not describe who responded and who did not respond. | 0, Unclear. Random sample of census but no information about any that did not respond | 0, No. No power calculations or justification | 1, Yes. Stool and urine samples submitted and screened prior to the study start. | 0, Unclear. No definition of a positive and no quality control measures. | 1, yes. The sonographer was unaware of the infection status of individual | 0, unclear. Used a non-validated protocol. No mention of the training of the sonographer | 1, Yes. Trainer sonographer, wasn’t aware of the infection status | 0, not applicable. No model to include data in and there is no follow up | 0, no. No covariate information was provided (no age/gender/location information) | 0, no. No model | 5 | M |
| **Study ID** | **D1** | **D2** | **D3** | **D4** | **D5** | **D6** | **D7** | **D8** | **D9** | **D10** | **D11** | **D12** | **D13** | **D14** | **Score** | **Risk of bias** |
| Hoffmann et al, 2001^43^ | 1, yes. Specific aim and analyses match what is reported. | 1, yes. Entire rice farmer village eligible between Jan-July 1994. | 0, no. Sampling method was not provided but it appears that a 'census' approach was used | 0, no. Does not provide statistics on differences response rate but provides the total number of participants that consented to both microscopy AND USS | 1, yes. Everyone in the village was eligible. | 0, no. No sample size justification provided | 1, yes. Exposure was measured before outcome. | 1, yes. Person excreting eggs is positive for *Schistosoma infection*. | N/A | 0, no. Does not explicitly say that two USS measurement were taken. | 0, unclear. Does not state who measured infection status | 1, yes. Missing but less than 20%. (18.9% of the village did not provide both stool sample and consent for USS) | 1, yes. Covariates defined. | 0, no. Descriptive statistics only | 7 | M |
| Kamdem et al, 2019^44^ | 1, yes. Specific aim and analyses match what is reported | 1, yes. Schoolchildren in rural, *S. mansoni* endemic Cameroon between Sept to Dec 2018. | 0, no. No details given about sampling method | 0, no. Differences in response rate not given nor were differences in characteristics | 1, yes. Clear eligibility criteria | 0, no. No sample size justification provided. | 0, unclear. Does not explicitly say the KK was done before USS | 1, yes. Both infection status and infection intensity were defined | N/A | 0, no. Does not explicitly say that two USS measurement were taken but they followed the Niamey protocol. | 1, yes. Microscopy = technicians, USS= clinicians | 0, no. 727/1002 consent children did not provide both parasitology and USS | 1, yes. Age groups were the main risk factor. | 0, no. Descriptive statistics only | 6 | M |
| Kariuki et al, 2001^28^ | 1, yes. Specific aim and analyses match what is reported. | 1, yes. Time period given (Oct 1998 to Jan 1999) and all >5 years in the study villages. | 0, no. Sampling method was not provided | 1, yes. 75.5% of eligible participants enrolled. | 1, yes. Clear eligibility criteria. | 0, no. No sample size justification provided | 0, unclear. Does not explicitly say the KK was done before USS | 1, yes. Infection status provided and infection intensity was handled as a continuous variable | 0, unclear. | 1, yes. More than two measurements taken. | 1, yes. Microscopy = experience technicians, USS= study authors | 0, no. 53% of the total participants were not in full analysis. | 1, yes | 0, no. Descriptive statistics only | 8 | M |
| **Study ID** | **D1** | **D2** | **D3** | **D4** | **D5** | **D6** | **D7** | **D8** | **D9** | **D10** | **D11** | **D12** | **D13** | **D14** | **Score** | **Risk of bias** |
| King et al, 2003^45^ | 1, yes. Specific aim and analyses match what is reported. | 1, yes. Two study populations defined in two setting countries | 0, no. Sampling method was not provided but ‘census’ approach taken. | 1, yes. No non-responders | 1, yes. Kenya- Everyone in the district was examined, 40% of the Egyptian village was examined. | 0, no. No sample size justification provided. | 0, unclear. Does not explicitly say the KK was done before USS | 1, yes. Infection status provided and infection intensity was handled as a continuous variable | N/A | 1, yes. More than two measurements taken. | 0, unclear. Does not state who measured infection status. | 1, yes. Missing data: Egypt 8.6%, Kenya 1% | 1, yes. | 0, no. Only adjusted covariates reported. | 8 | M |
| Mazigo et al, 2015^46^ | 1, yes. Specific aim and analyses match what is reported. | 1, yes. Yes, permanent resident of study area. | 1, yes. Described in the 'base' study. | 0, no. No information given about non-responders or differences between known characteristics | 1, yes. Clear inclusion and exclusion criteria | 1, yes. Sample size calculation is given in referenced paper. | 1, yes. Exposure was measured before outcome. | 1, yes. Infection status and intensity defined. 10% of the samples were quality assessed | N/A | 1, yes. Six measurements were taken | 1, yes. Microscopy = technicians, USS= radiographer. Blinded. | 1, yes. No missing data. | 1, yes. All covariates listed and defined. | 1, yes. Both unadjusted and adjusted effect measures are reported. | 12 | L |
| Meurs et al, 2012^47^ | 1, yes. Specific aim and analyses match what is reported. | 0, no. Does not describe the exact study population | 0, no. No method of selection described | 0, no. Does not describe who responded and who did not respond. | 1, yes. Eligibility criteria provided and specific time frame given (July to Nov 2009). | 0, no. No sample size justification provided | 0, unclear. Does not explicitly say the KK was done before USS | 1, yes. | 1, yes. Technicians were blinded | 0, no. Only one measurement taken (Image pattern). | 1, yes. Clinicians who conducted USS were blind to infection status of patients. | 1, yes. No missing data. | 1, yes. All covariates listed and defined. | 1, yes. Both unadjusted and adjusted effect measures are reported. | 8 | M |
| **Study ID** | **D1** | **D2** | **D3** | **D4** | **D5** | **D6** | **D7** | **D8** | **D9** | **D10** | **D11** | **D12** | **D13** | **D14** | **Score** | **Risk of bias** |
| Mohammed-Ali 1999 | 1, yes clear aims | 0, No. Detailed information about the population but no study year | 1, yes. Everyone in the study area included apart from those unavailable at the time of ultrasound | 1, yes. 88% of the population were involved in the study and details on the wider community. | 1, yes. Census approach to sampling | 0, no. No power calculations or justification | 1, yes. Stool and urine samples submitted and screened prior to the study start. | 0, unclear. No clear definition or quality control measures or who conducted the microscopy. | 0, unclear. No mention of who performed KK/USS and their knowledge of the status | 0, no. No validated methodology, but trained sonographer and more than 2 measurements | 0, unclear. No mention of who conducted the microscopy or the sonography. | 0, not applicable. No model to include data in and there is no follow up | 1, yes. Clear age groups | 0, No. No model | 6 | M |
| Motawea et al, 2004^48^ | 1, yes. Specific aim and analyses match what is reported. | 0, no.  Time period not specified. | 1, yes. Two stage study. Systematic random sample of Ezbas and mother village. | 0, no. No information given about non-responders or differences between known characteristics | 1, yes. Clear eligibility criteria. | 0, no. No sample size justification provided | 0, unclear. Does not explicitly say the KK was done before USS | 0, no. Does not explicitly define what infection status or intensity is, even looking at the reference to the technique in the full text | 0, unclear.  Does not report whether USS assessor were aware of PPF status of cases/controls. | 0, no. Does not mention who measured the outcome | 0, unclear. Does not state who measured infection status or PPF | 0, unclear. Does not report who is missing from the study | 1, yes. No missing data from case-control part of study. | 0, no. Descriptive statistics only. | 4 | H |
| Mueller et al, 2019^31^ | 1, yes. Specific aim and analyses match what is reported | 1, yes. Study population clearly defined. | 1, yes. All individuals above 1 year. | 1, yes. Everyone took part in the study. | 1, yes. All the study participants  in Magu district took part | 1, yes. Sample size calculation provided. | 0, unclear. Does not explicitly say the KK was done before USS | 1, yes. Infection intensity defined and categorised into WHO intensity criteria. 15% of samples re-checked for quality. | N/A | 1, yes. Multiple measurements taken | 1, yes. Microscopy = technicians, USS= sonographers | 1, yes. No missing data. | 1, yes. All covariates listed and defined. | 1, yes. Both unadjusted and adjusted effect measures are reported | 12 | L |
| **Study ID** | **D1** | **D2** | **D3** | **D4** | **D5** | **D6** | **D7** | **D8** | **D9** | **D10** | **D11** | **D12** | **D13** | **D14** | **Score** | **Risk of bias** |
| Nalugwa et al, 2017^49^ | 1, yes. Specific aim and analyses match what is reported | 1, yes. Children in *S. mansoni* endemic communities during April 2013- Feb 2014 | 1, yes. Convenience sampling | 0, no. Does not provide statistics on response rate. | 0, unclear. The fishing communities were randomly selected but the participants were then sampled out of convenience | 0, no. No sample size justification provided | 1, yes. Exposure was measured before outcome | 1, yes. Clearly defined exposure and 10% of the samples were re-checked for quality assurance | N/A | 0, no. Only one measurement taken for PPF. (Image pattern) | 1, yes. Microscopy = microscopist, USS= sonographers | 0, unclear. Does not report whether there is missing data. | 1, yes. All covariates defined. | 0, no. Descriptive statistics only | 7 | M |
| Ndamba et al, 1991 | 1. yes. Specific aim and analyses match what is reported | 0, no. No study year | 0, no. Insufficient details of how the cases and controls were selected. | 1, yes. 91% of the study population were examined. | 1, yes. Cases and controls were from the same population and matched for age. | 0, no. No power calculations or justification | 1, yes. Stool samples were collected before the ultrasonography was obtained | 0, unclear. No definition of a positive case and no quality control measures. | 1, yes. The ultrasound was unaware of the infection status of individual | 0, unclear. Doesn’t state the trained status of the sonographer | 1. yes. Read by three technicians and a sperate person for sonography | 0, not applicable. No model to include data in and there is no follow up | 1, yes. Covariates for egg intensity and age presented | 0, No. No model | 7 | L |
| Nega et al, 2014^50^ | 1, yes. Specific aim and analyses match what is reported | 0, no. No time period provided | 1, yes. Sample size calculation provided | 0, no. Response rate was not given nor where the differences in characteristics statistically assessed. | 0, no. No eligibility criteria provided | 1, yes. Sample size calculation is given. | 0, unclear. Does not explicitly say the KK was done before USS | 1, yes. Infection intensity defined and categorised into- light, moderate and heavy. | N/A | 0, no. Does not explicitly say that two USS measurement were taken | 0, unclear. Does not state who measured infection status or PPF | 1, yes. No missing data. | 0, no. The age of youngest participant is not given, only (‘<14’ for the lowest age category) | 0, no. Descriptive statistics only and logistic regression outputs of were not provided, only p-values | 5 | M |
| **Study ID** | **D1** | **D2** | **D3** | **D4** | **D5** | **D6** | **D7** | **D8** | **D9** | **D10** | **D11** | **D12** | **D13** | **D14** | **Score** | **Risk of bias** |
| Negrao-Correa et al, 2014^29^ | 1, yes. Specific aim and analyses match what is reported | 0, no.  Time period not specified . | 0, no. Does not report sampling strategy | 0, no. Response rate was not given nor where the differences in characteristics statistically assessed. | 0, unclear. As a of unreported sampling method despite eligibility criteria being provided | 0, no. No sample size justification provided | 0, unclear. Does not explicitly say the KK was done before USS | 0, no. Exposure definitions not reported | N/A | 1, yes. Multiple measurements taken | 0, unclear. Does not state who measured infection status | 1, yes. No missing data. | 1, yes. All covariates defined. | 0, no. Descriptive statistics only | 4 | H |
| Nooman et al, 1995^51^ | 1, yes. Specific aim and analyses match what is reported | 1, yes. Protocol for study described the study population of interest. | 1, yes. Stratified random sampling applied | 1, yes. Microscopy sample (36.0%).Response rates in USS (91.0%) | 1, yes. Clear inclusion criteria and to be representative of governorate’s rural population. | 1, yes. Justification of sample size given in study design paper | 0, unclear. Does not explicitly say the KK was done before USS | 1, yes. Infection intensity defined and standardised microscopy measurement techniques. | N/A | 0, no. Does not explicitly say that two USS measurement were taken | 1, yes. Microscopy = technicians, USS= clinicians | 0, unclear. Does not report who is missing from the study | 1, yes. Risk factors defined and categorised. | 0, unclear. Does not describe how the effect measures were obtained | 9 | M |
| Nooman et al, 2000^52^ | 1, yes. Specific aim and analyses match what is reported | 0, no.  Time period not specified . | 1, yes. Multi-stage, systematic random sampling | 0, no. Response rate was not given nor where the differences in characteristics statistically assessed. | 1, yes. Broad eligibility criteria | 0, no. No sample size justification given. | 0, unclear. Does not explicitly say the KK was done before USS | 0, no. Does not explicitly define what infection status is, even looking at the reference to the technique in the full text | N/A | 1, yes. Three measurements taken | 1, yes. Microscopy = technicians, USS= sonographers | 0, no. Only 20% of the study population underwent USS | 1, yes. All covariates defined. | 0, no. Descriptive statistics only | 6 | M |
| **Study ID** | **D1** | **D2** | **D3** | **D4** | **D5** | **D6** | **D7** | **D8** | **D9** | **D10** | **D11** | **D12** | **D13** | **D14** | **Score** | **Risk of bias** |
| Ocama et al, 2017^53^ | 1, yes. Specific aim and analyses match what is reported | 0, no. No time period specified. | 1, yes. First 20 patients that arrive each day at the clinic. | 0, no. No information given about non-responders or differences between known characteristics | 0, no. People attending the hospital clinic might not be representative of all adults over 18. | 0, no. No sample size justification provided | 1, yes. Exposure was measured before outcome. | 0, unclear. Commercial diagnostic used but not clear whether test was conducted in a reference laboratory. | N/A | 0, no. Does not explicitly say that two USS measurement were taken | 1, yes. Microscopy = trained laboratory technician, USS = trained radiographer | 0, unclear. | 1, yes. No missing data | 0, no. Covariates not applied consistently. Focuses on PPF only in the 30-50 years age category. | 5 | M |
| Odongo-Aginya et al, 2010^30^ | 1, yes. Specific aim and analyses match what is reported | 1, yes. Clear description of study population provided | 0, no. No details about sampling method | 0, no. No information given about non-responders or differences between known characteristics | 1, yes. Clear inclusion and exclusion criteria | 0, no. No sample size justification provided | 1, yes. USS examiners were blinded to the patients' infection status and conducted exams following KK | 1, yes. Infection intensity defined and categorised into- light, moderate and heavy, although not using standard WHO categories | N/A | 1, yes. Three measurements were taken. | 0, unclear. Does not state who measured infection status or PPF | 0, unclear. Does not report who is missing from the study. | 0, no. Age category is not consistent and skips an entire section (21-29) | 0, no. Descriptive statistics only | 6 | M |
| Oliveira et al, 2006^54^ | 1, yes. Specific aim and analyses match what is reported. | 0, no.  Time period not specified nor eligible individuals | 0, no. No details about sampling method. | 0, no. No information given about non-responders or differences between known characteristics | 0, unclear. Cannot follow the rationale for the 91 participants from the 644 residents in the study area | 0, no. No sample size justification provided | 0, unclear. Does not explicitly say the KK was done before USS | 1, yes. Infection intensity described but non-standard categorisation (0 EPG, 12-100 EPG and >100 EPG) | N/A | 0, no. Does not explicitly say that two USS measurement were taken. | 0, unclear. Does not state who measured infection status | 0, unclear. Does not report who is missing from the study | 0, no. No justifications given for the categorisation of the covariates. | 0, no. Descriptive statistics only | 2 | H |
| **Study ID** | **D1** | **D2** | **D3** | **D4** | **D5** | **D6** | **D7** | **D8** | **D9** | **D10** | **D11** | **D12** | **D13** | **D14** | **Score** | **Risk of bias** |
| Olveda et al, 2017^55^ | 1, yes. Specific aim and analyses match what is reported | 1, yes. Flowchart provided explaining the demographics of the included participants | 0, no. No sampling method provided. | 0, no. No information given about non-responders or differences between known characteristics | 0, unclear. The sample is not randomly selected, and the villagers could opt not to take part | 0, no. No sample size justification provided | 0, unclear. Does not explicitly say the KK was done before USS | 1, yes. Both exposure defined. | N/A | 0, no. Does not explicitly say that two USS measurements were taken | 1, yes. Microscopy = technicians, USS= sonographers | 0, no. Only 151/565 (26.7%) of study participants provided both infection intensity and PPF grade | 1, yes. Covariates defined. | 0, no. Descriptive statistics only | 5 | M |
| Roquet et al, 1993 | 1, yes. Specific aim and analyses match what is reported | 1, yes. Noted location, inclusion criteria, demographic table and study year | 1, yes. Radom sampling, then selected participants positive for schistosomiasis | 1, yes 70% responded to invitation for further study after initial parasitological survey. | 1, yes. Controls were matches of age and ethnic group but they had to be sampled from an alternative area | 0, No. No power calculations or justification | 1, Yes. Infection status determined before the sonography | 0, Unlcear. No clear definition or quality control measures. Measured unsing the modified KK technician | 0, unclear. No mention of who performed KK/USS and their knowledge of the status | 0, No. No validated methodology, but trained sonographer and more than 2 measurements | 0, unlcear. No mention of who conducted the microscopy but there was a trained sonographer | 0, not applicable no model | 1, yes. Clear age groups | 0, No. No model | 7 | M |
| Ruiz et al, 2002^56^ | 1, yes. Specific aim and analyses match what is reported | 0, no. No time period provided | 0, no. No details about sampling method | 0, no. No information given about non-responders or differences between known characteristics | 0, no. No clear eligibility  criteria. | 0, no. No sample size justification provided | 0, unclear. Does not explicitly say the KK was done before USS | 1, yes. Exposures defined. | N/A | 0, unclear. Seven USS measurements were taken using Cairo protocol. | 0, unclear. Does not state who measured infection status. | 1, yes. No missing data. | 0, no. Only the exposure and outcomes are defined | 0, no. Descriptive statistics only | 3 | H |
| **Study ID** | **D1** | **D2** | **D3** | **D4** | **D5** | **D6** | **D7** | **D8** | **D9** | **D10** | **D11** | **D12** | **D13** | **D14** | **Score** | **Risk of bias** |
| Russell et al, 2020^57^ | 1, yes. Specific aim and analyses match what is reported | 1, yes. Time-period provided, study location and also the demographics of the study population. | 1, yes. Stratified random sampling. | 0, no. No information given about non-responders. | 1, yes. Children were randomly from the school register. | 1, yes. Maximum 300 in the sample due to resource scarcity | 0, unclear. Does not explicitly say the KK was done before USS | 1, yes | N/A | 0, no. Only one measurement taken (Image pattern) | 1, microscopy = technicians, USS= trained medical students | 1, yes. Only 8.3% participants were missing data. | 1, yes. All covariates were defined. | 1, yes. Both adjusted and unadjusted odds ratios presented. | 10 | L |
| Sayasone et al, 2012^58^ | 1, yes. Specific aim and analyses match what is reported | 1, yes. Time-period provided, study location and also the demographics of the study population. | 0, no. No details about sampling method. | 0, no. No information given about non-responders or differences between known characteristics | 0, unclear. There might be differences between people who submit a stool sample and those who do not | 0, no. No sample size justification provided | 0, unclear. Does not explicitly say the KK was done before USS. Blind | 0, no. Does not explicitly define what infection status or intensity is, even looking at the reference to the technique in the full text | N/A | 1, yes. Multiple measurements taken following Cairo protocol. | 0, unclear. Does not state who measured infection status | 0, unclear. Does not report who is missing from the study | 1, yes | 0, unclear. Not reported whether the effect measures were adjusted/unadjusted | 4 | H |
| Silveira et al, 2002^59^ | 1, yes. Specific aim and analyses match what is reported | 0, no. No study time period reported | 0, no. No sampling strategy reported | 0, no. No information given about non-responders or differences between known characteristics | 0, unclear. No rationale provided for the participants included | 0, no. No sample size justification provided | 1, yes. Stool samples examined in May 1998 and USS completed in Sept 1998 | 1, yes. Exposures are defined. | N/A | 1, yes. Multiple measurements taken using the Cairo protocol. | 0, unclear. Does not state who measured infection status or PPF | 1, yes. No missing data. | 1, yes. Clearly defined covariates in statistics section. | 0, no. Descriptive statistics only | 6 | M |
| Wiegand et al, 2021^60^ | 1, yes. Specific aim and analyses match what is reported | 1, yes. Clearly describes each study country’s study populations in the supplementary material. | 1, yes. Multi-stage random sampling. | 0, no. No information given about non-responders or differences between known characteristics | 1, yes. | 0, no. No sample size justification provided | 0, unclear. Does not explicitly say the KK was done before USS | 1, yes. Infection status defined as the presence of eggs in stool. Infection intensity categorised using WHO categories. | N/A | 1, yes. At least two measurements were taken (Image Pattern, portal vein diameter) | 0, unclear. Does not state who measured infection status or PPF | 0, unclear. Study does not report missing data | 1, yes. All covariates in the study were defined. | 0, unclear. Not reported whether the effect measures were adjusted/unadjusted | 7 | M |

*L –Low risk of bias*

*M- Medium risk of bias*

*H- high risk of bias*

Figure S2: Publication bias as shown by funnel plot

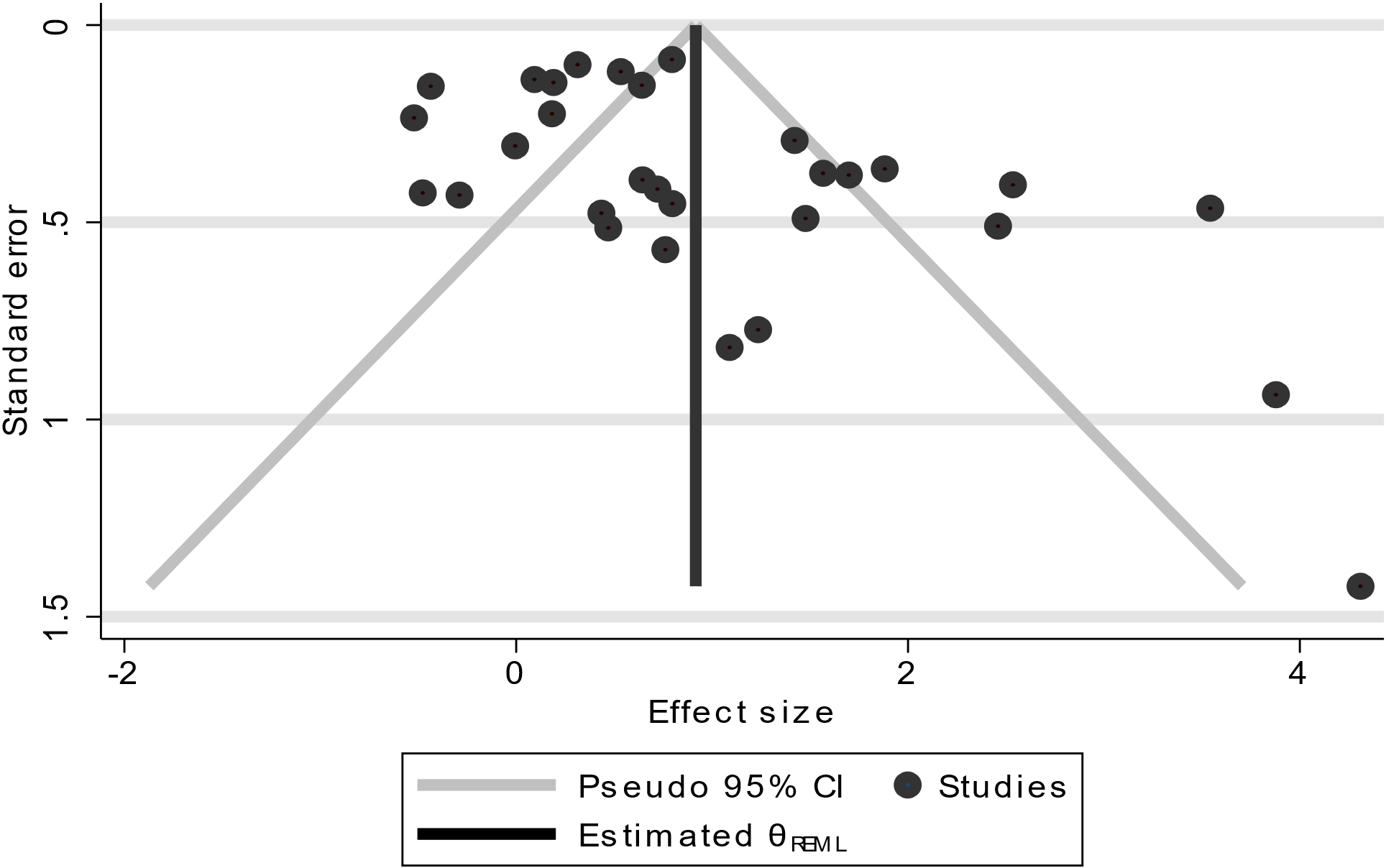
